# Personalized nutrition products in a hospital setting: A prospective proof-of-concept study of acceptability in hospitalized patients and staff

**DOI:** 10.64898/2026.10.01.26364528

**Authors:** M Melchers, S van der Haar, FPM Hoevenaars, S Wopereis, S van Beek, M Ummels, K van Bokhoven, M Hoppenbrouwers, A Rijfers, M Menkveld, MWJ Noort, ARH van Zanten, IWK Kouw

## Abstract

**Background & Aims:** Personalized nutrition has the potential to improve nutritional intake and clinical outcomes in hospitalized patients by adapting to individual dietary needs and preferences. Despite its promise, no personalized nutrition product (PNP) for hospitalized patients is currently available. This study aimed to assess the acceptability of PNPs among hospitalized patients and hospital staff.

**Methods:** A prospective, ten-days, proof-of-concept study was conducted among in-hospital adult patients (≥ 18y). Study patients received a freshly 3D printed PNP, customized to match individual taste preferences and health profiles. Patients could reorder the product daily if they remained hospitalized >24h after consumption. Consumption was assessed by weighing the PNP before and after consumption. Acceptability, sensory feedback, and overall experience were evaluated using questionnaires. Hospital staff also evaluated sensory feedback and acceptability. Factors influencing consumption and liking among patients were evaluated by testing within-group differences. Correlations between consumption rates, liking and patient characteristics were assessed.

**Results:** Eighty-seven patients received 137 PNPs during the study period. The average overall liking score was 5.7 (±2.1) on a 1 – 9 scale, and median consumption rate was 51% [Interquartile Range 19 – 100]. Consumption rates positively correlated with overall liking (R^2^ = 0.50, *p* < 0.001). Patients’ willingness to reorder was 3.5 (±2.3) on a 1–7 scale, and reorder rate 63%. The perceived value of a PNP service in a hospital setting (scale 1 – 7) was scored 5.3 (±1.7) by patients and 4.5 (±1.1) by staff.

**Conclusions:** PNPs were evaluated as moderately positive in terms of acceptability and value by both patients and staff. However, further optimization of the product is needed before evaluating clinical efficacy.

## Background

Malnutrition is highly prevalent in hospitalized patients, with ∼20-50% of patients being at-risk of malnutrition during hospitalization, a condition that is associated with increased length of hospital stay, and increased mortality and morbidity (1, 2). Hospitalized patients at risk of malnutrition often have food intakes 30-40% below the recommended levels (2, 3). Altered appetite and changes in taste are commonly reported by hospitalized patients, as a result of inflammation, medication side effects, certain comorbidities and ageing (4, 5). These nutrition-impacting symptoms are suggested to affect nutrition intake (6), and are related to poor patient-related outcome (4).

The concept of personalized nutrition (PN) considers individual nutritional requirements as well as factors like food preferences. It utilises individual genetic, biochemical, and phenotypic characteristics to guide tailored dietary recommendations for optimizing health outcomes and managing diseases (7). Individualized nutritional support tailored to individual preferences and dietary needs has shown to improve nutritional intake of specific nutrients, and consequently clinical outcomes of hospitalized patients (1). The current European Society for Clinical Nutrition and Metabolism (ESPEN) guidelines for hospitalized patients emphasize the importance of personalized and adaptive nutritional care strategies tailored to the patient’s specific condition, dietary needs, and food preferences (8). Several individualized nutrition strategies in clinical practise are available, including digital information systems providing patients assessments on biochemical, phenotypic, and lifestyle parameters to assist on personalized medical strategies (9), and routine use of bed-side energy expenditure measurements to establish PN therapy recommendations in critical care settings (10). However, a ready-to-eat personalized nutritional product (PNP) to meet a patient’s food preferences and personalized nutritional needs has been unexplored in medical settings.

A flexible food manufacturing platform (FFMP) utilizing innovative technologies, including three-dimensional (3D) food printing, may provide patients an on-demand, ready-to-eat PNP based on their individual daily needs, taste, and texture preferences. Patients with specific medical conditions can benefit from personalized meal options tailored to their dietary needs and treatment plans (11). However, acceptance of new food technologies and products can be challenging due to its newness and unfamiliarity among consumers (12). Little is known about the experiences and acceptance of a PNP among patients and hospital staff, which currently hinders further exploration of these innovative products’ health benefits in a medical setting on a large scale. Additionally, a better understanding of patient characteristics and food preferences and liking will inform further research on the efficacy of a PN service in a hospital setting.

This proof-of-concept study aimed to evaluate patient and staff acceptance of a 3D-printed PNP tailored to taste preference and medical condition in a hospital setting. We investigated product’s sensory liking and consumption rates, and attitudes towards the concept among hospitalized patients who received a PNP. Additionally, sensory liking of these products and acceptance of PN service were evaluated in hospital staff involved in nutrition care.

## Material & methods

### Study design

A single-center, prospective, proof-of-concept study in hospitalized patients and staff was conducted in a teaching-affiliated hospital in Ede, the Netherlands, between June 23 and July 5, 2024. As this was a proof-of-concept study, no formal sample size calculation was performed. The study aimed to include all eligible patients admitted during the predefined two-week study period who met the inclusion criteria and provided informed consent. The sample size was considered sufficient to evaluate the feasibility and acceptability of the PNP and provide estimates to inform the design of future studies.

### Ethical Procedures

All study patients were informed on the purpose of the study and were able to consider participation in the study. Patients were not informed about the production process of the PNP to minimize the risk of bias (incomplete disclosure) since we observed that hospitalized patients report both positive and negative associations towards the idea of 3D-printed PNPs in earlier work conducted in the same hospital (*unpublished data*). Following completion of the study, all participants were informed about the production technique of the PNP. All eligible participants provided written informed consent before participation in the study. A declaration confirming that the study did not fall under the Medical Research Involving Human Subjects Act was obtained (NR 2023-64). Subsequently, the research protocol was approved by the local ethical committee of Hospital Gelderse Vallei, Ede, the Netherlands (NR 2403-020). The study was conducted in accordance with the ethical standards outlined in the World Medical Association of Helsinki Declaration of 2013. Participants’ data were collected under the Personal Data Protection Act and used pseudo-anonymized. The study was reported in accordance with the STROBE guidelines (13).

### Personalized Nutrition Product

#### Production

This study was part of a larger project with the aim to develop and test a complete PN service for designing, producing, and distributing freshly 3D-printed PNPs based on personal nutritional needs and taste preference (Nutri-Bites®, Noort, M. *et al*. (2026). *Manufacturing Personalized Nutrition Food products using a flexible food production platform, unpublished manuscript*). The PN service was also tested in a ten-days proof-of-concept study in a military setting (van der Haar, S. *et al*. (2026). *Consumer acceptance of Personalized Nutrition Products in a Real-Life Military Setting: a Proof-of-Concept Study*, *unpublished manuscript*). In short, the PNPs (example shown in **Figure 1**) were produced using a for this purpose developed FFMP. The FFMP is a fully automated mini food-factory, combining 3D food printing with smart dosing to compose individual food products. The extrusion based 3D printing technique involves layer-by-layer deposition dough or paste-like food materials to produce a pre-designed digital 3D model (14). The FFMP contains several liquid- and powder deposition stations for the incorporation of a diverse range of micronutrient powders, fillings, and toppings based on a patient’s medical conditions and anticipated health benefits.

**Figure 1.**
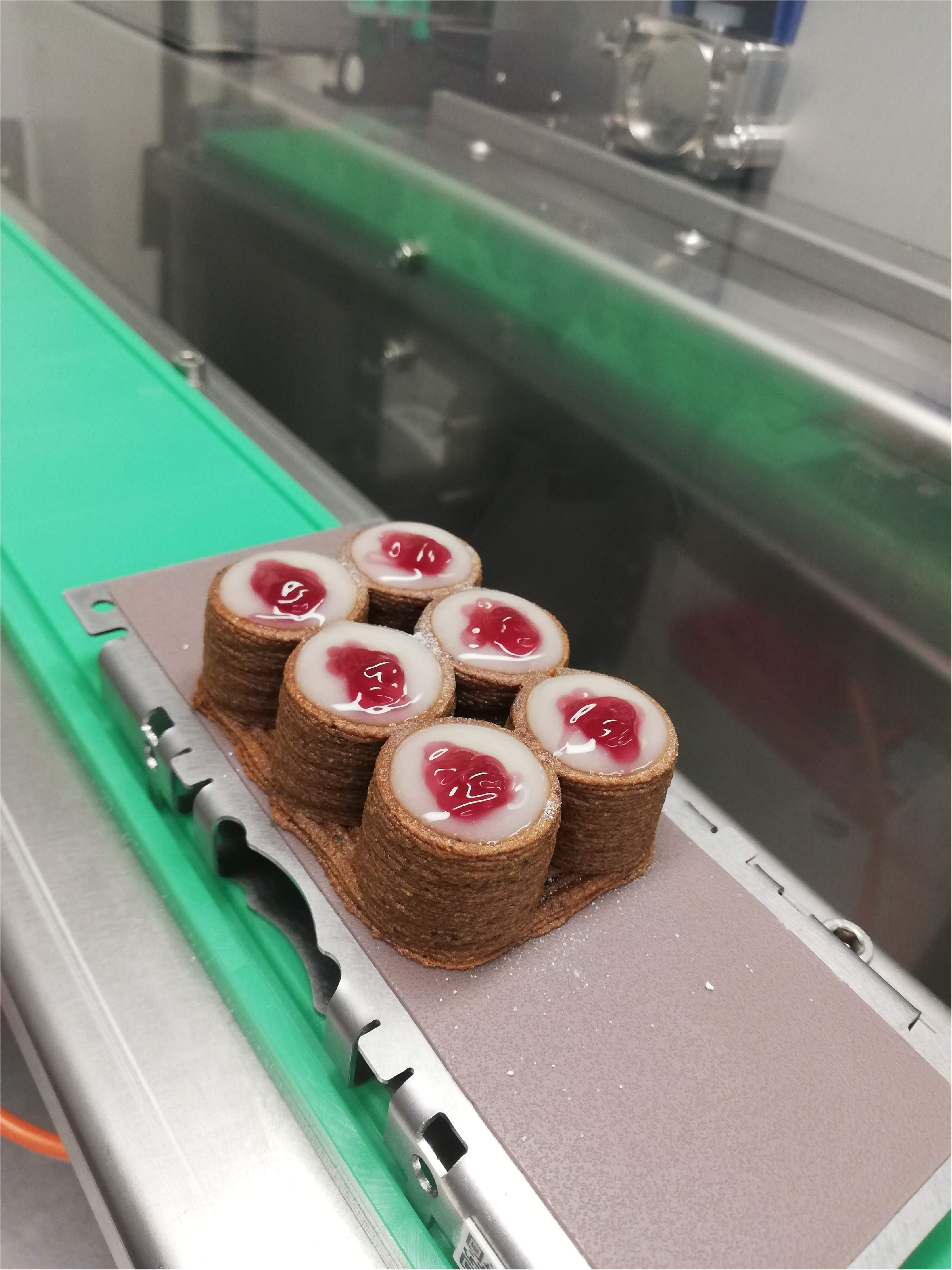
A Personalized Nutrition Product as produced in the FFMP. In this example the PNP is composed of a plum-based cellular casing, a micronutrient powder, a yoghurt filling, and summer-fruit topping.

#### Composition

Six different pre-defined health profiles were created based upon prevalent medical conditions in hospitalized patients and an anticipated health benefit: 1) muscle health, 2) immune health, 3) blood glucose regulating, 4) energy restricted, 5) cognition, and 6) wellbeing. Health nutritional profiles were formulated upon extensive literature review. In brief, first the European register of health claims was queried for nutrition claims concerning the chosen health benefits. Next, a health taxonomy was used in combination with a text mining strategy to query for nutrients with potential health outcomes in PubMed (National Center for Biotechnology Information (NCBI). National Library of Medicine, USA). Outcomes were evaluated based upon evidence quality and product formulation demands which resulted in defined macronutrient ranges and micronutrient contents per health profile (Supplemental Table 1a). During the study, a physician and dietitian involved in the study (MM and SvB) decided on the health benefit most relevant to the patient, whereas the patient selected the preferred flavour for the product. This determined the type of casing, type of filling, and the size of the PNP to be produced. The composition of the PNP are presented in Supplemental Table 1b.The filling and the casing dough of the PNP was health profile-dependent and contained specific added micronutrients and amino acids, not exceeding the upper tolerable limits. Each PNP was topped off with a flavoured topping of choice (lemon, orange, chocolate, caramel, strawberry, or summer fruit). The products weighed between 50 – 100 g, dependent on health profile. Extensive motivation for the product’s compositions and fillings is discussed in a separate paper (Noort, M. *et al*. (2026). *Manufacturing Personalized Nutrition Food products using a flexible food production platform, unpublished manuscript)*

### Study patients

#### Population

Adult (≥18y) patients admitted to the internal medicine, neurology, orthopaedic surgery, pulmonology, gastroenterology, cardiology, or general surgical ward with an expected hospital stay of at least 48h were eligible for participation. Exclusion criteria for participants were: 1) any known food intolerances or allergies, 2) deemed incompetent to (safely) consume the PNP (e.g. swallowing difficulties, too ill, patient had to remain in a fasting state for the purpose of diagnostics or intervention) by the treating physician and/or study physician, and 3) deemed incompetent to provide well-considered informed consent (e.g. cognitive problems, delirium, or mental disorders) by the treating physician and/or study physician.

#### Study procedures

After obtaining informed consent, a PNP health profile was selected by the study physician (MM) and dietitian (SvB) based on relevant personal information collected from the electronic patient database (Nexus, NEXT Generation, 2024), including relevant medical history, reason of hospital admission, medication use, and risk of malnutrition (assessed by Short Nutritional Assessment Questionnaire (SNAQ) score). Patients were able to customize their PNP by selecting the flavour of the filling and topping through a digital ordering platform on a digital tablet (Diamonds, TNO, the Netherlands) that was connected to the FFMP that was located at Netherlands Organization for Applied Scientific Research (TNO, Eindhoven, the Netherlands). The PNP was produced the following morning between 08:00 AM and 12:00 PM and transported to the hospital in cool boxes. All products were weighed and distributed among the study patients in the afternoon around 03:00 PM as in-between meal snack. Production and distribution of the PNP was performed only during weekdays during the study period. A short description regarding the selected health profile was handed out at distribution. Patients were instructed upon distribution to consume the product themselves and to complete a paper questionnaire on product liking, appearance, and sensory perceptions during consumption of the PNP. One hour after distribution, these scores were filled out in an electronic questionnaire (EyeQuestion Software, Logic 8) on a digital tablet, alongside additional questions regarding patient experiences, willingness to reorder, reason to reorder or not to reorder, and acceptability of the concept (Supplemental File 1). Although there was some variation between the time of production and consumption, all products were consumed within eight hours after production.

### Data collection

#### Patient’s characteristics

Characteristics and demographics collected from the electronic patient medical records (Nexus, NEXT Generation, 2024) were: age, sex, body weight, height, body mass index (BMI), smoking status, alcohol use, relevant medication use, and comorbidities included in the updated Charlson Comorbidity Index (CCI) (15). Baseline laboratory values (C-reactive protein (CRP), white blood cell count (WBC), and albumin blood levels) were collected within 72h of the first product distribution. Nutritional status characteristics that were collected included the SNAQ score, prescription of energy- and protein-enriched diet, and receiving nutritional support (parenteral nutrition, enteral tube feeding, and/or oral nutritional supplements (ONS)).

#### Sensory evaluation

The degree of sensory liking of the PNP was assessed using the 9-point Hedonic Scale, which is extensively used in sensory evaluations, and ranges from ‘dislike extremely’ (=1) to ‘like extremely’ (=9). Each product was rated on the attributes of overall liking, taste, aftertaste, texture, appearance, and freshness through a short paper questionnaire; patients were instructed to complete it during consumption. Any report of physical complaints was asked as an open question one hour after consumption.

#### Product consumption

Product consumption rates (g and % of provided product) were quantified by weighing the PNP before and after consumption using a digital platform scale (Steward Systems ®). Protein and energy intake per consumed PNP were calculated based on the product composition (Supplemental Table 1b) and total amount consumed.

#### Reordering

One hour after consumption, the degree of willingness to reorder a PNP the following day was assessed using a 7-point Hedonic scale. Eligibility to reorder the following day (i.e. expected to remain in the hospital for another 24h, no restrictions for oral intake) was assessed through the electronic patient medical records and consultation of the attending physician. Study procedures were repeated in case an eligible patient reordered for the following day. Reasons for intentions to reorder or to not reorder were asked as an open question and were evaluated independent of eligibility to reorder.

#### Attitude and perceptions

Attitude and perceptions towards the PNP and the PN service in the hospital were assessed at the end of the study participation. Study patients who were ineligible to reorder or declined participation for the next day, completed additional questions regarding the acceptability and attitude towards the concept of PNP and its service on a 7-point agreement scale. Patients were then informed on the production process of the PNP by a 3D food printer and their awareness of the 3D printing technique used for the production of the PNP was asked.

### Staff

#### Respondents and study procedures

Hospital employees involved in the development and management of the hospital food service system and day-to-day nutritional care were invited to voluntarily participate in a one-hour session on the final day of the ten-days study period. During this session information on the PNP service was provided through a video illustrating the process of PNP ordering, production by the FFMP, and distribution among patients was shown. Furthermore, additional information was given regarding product composition and participants were able to taste a PNP (randomized composition of health profile, filling, and topping). All staff completed an electronic questionnaire on product liking (Hedonic Scale 1 – 9) and their opinion regarding acceptability, perceived values and benefits, intention to adopt and implement, and practical feasibility in a hospital setting (agreement scale 1 – 7).

### Statistical analysis

All scores on Likert scales are reported as mean ± standard deviation (SD) independent of distribution. Other continuous variables are described as median alongside interquartile ranges [IQR] or mean alongside SD in case of normally distributed data. Normality was tested by inspection of Q-Q plots and histograms. Differences in consumption rates and sensory evaluation within study patients were tested using Mann-Whitney U or Student’s T-test based on distribution. For differences in consumption and sensory evidence-based nutrition-impacting factors, Levene’s test was used to assess the equality of variances between groups. Bivariate correlations between consumption rates and patient characteristics are reported using Pearson correlations coefficients. Characteristics of interest included nutrition-impacting factors such as age, BMI, active cancer, certain medication use, smoking, alcohol use, nutritional status (SNAQ-score), and inflammatory parameters (i.e. CRP, WBC, and albumin levels). For within-group comparisons in patients that consumed and filled in a questionnaire more than one study day, consumption (rates) and sensory liking scores on the first study day were used. Repeated product distributions were included only in descriptive analyses of product consumption and reordering. Missing questionnaire data were not imputed. Analyses of sensory evaluation, attitudes, and reordering intentions were performed using available-case analysis, including only completed questionnaire for the respective outcome. Statistical significance was set at *p*<0.05. All statistical analyses were conducted using IBM SPSS Statistics 29.0 (IBM Corporation, Armonk, NY, USA) and graphs created using Prism (GraphPad Software, version 8.2.0).

## Results

### Patients

#### Study population

In total, 87 study patients received a PNP (Figure 2). Clinical characteristics of the study patients are displayed in Table 1. Median age was 69 y [IQR 61 – 78] and BMI was 26.0 kg/m^2^ [IQR 23.1 – 28.3]. The majority of patients was admitted to the internal medicine ward (25%), followed by surgical (20%), and pulmonology ward (18%).

**Figure 2:**
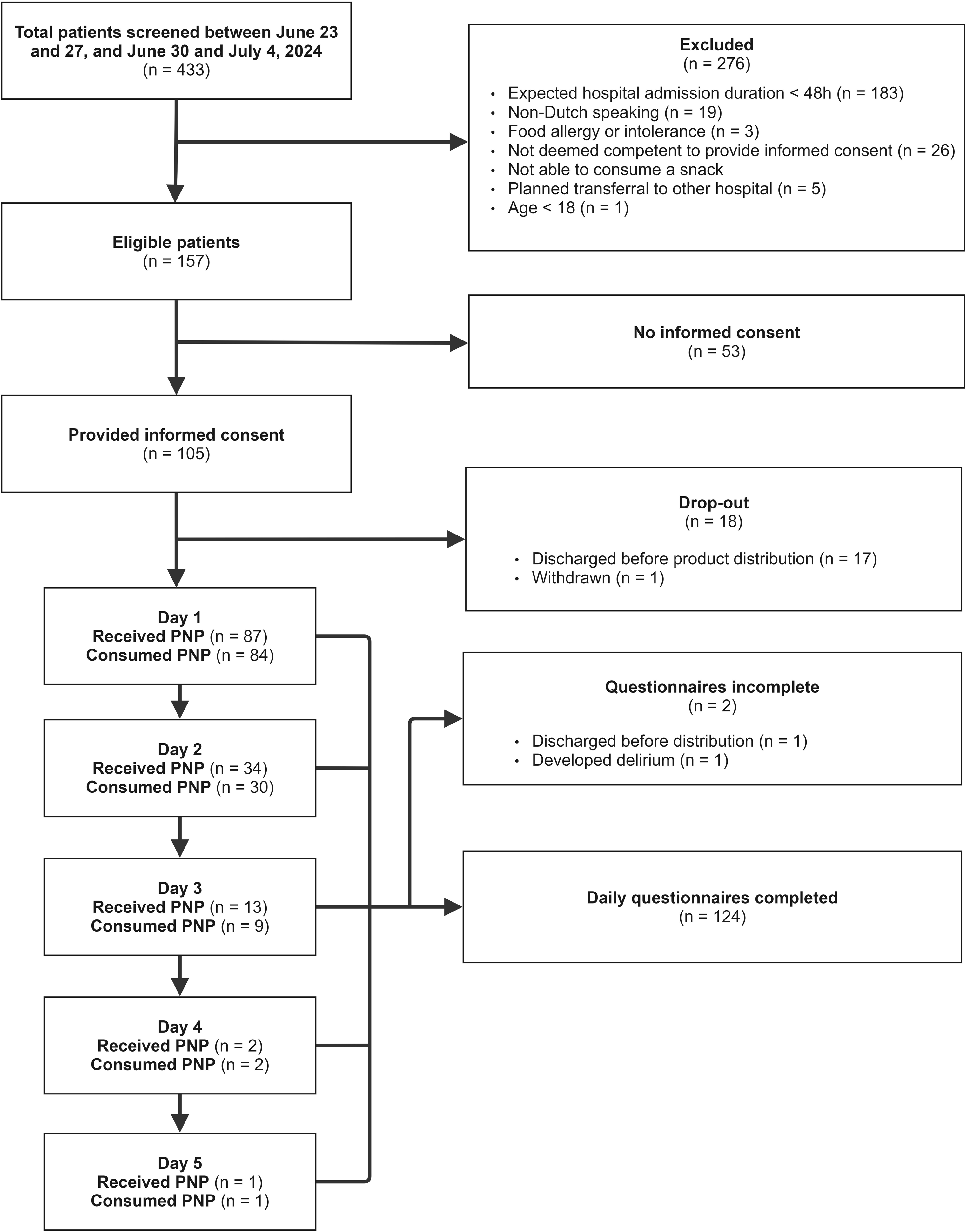
Flowchart displaying screening and recruitment of study patients and PNP distribution among included patients.

**Table 1:**
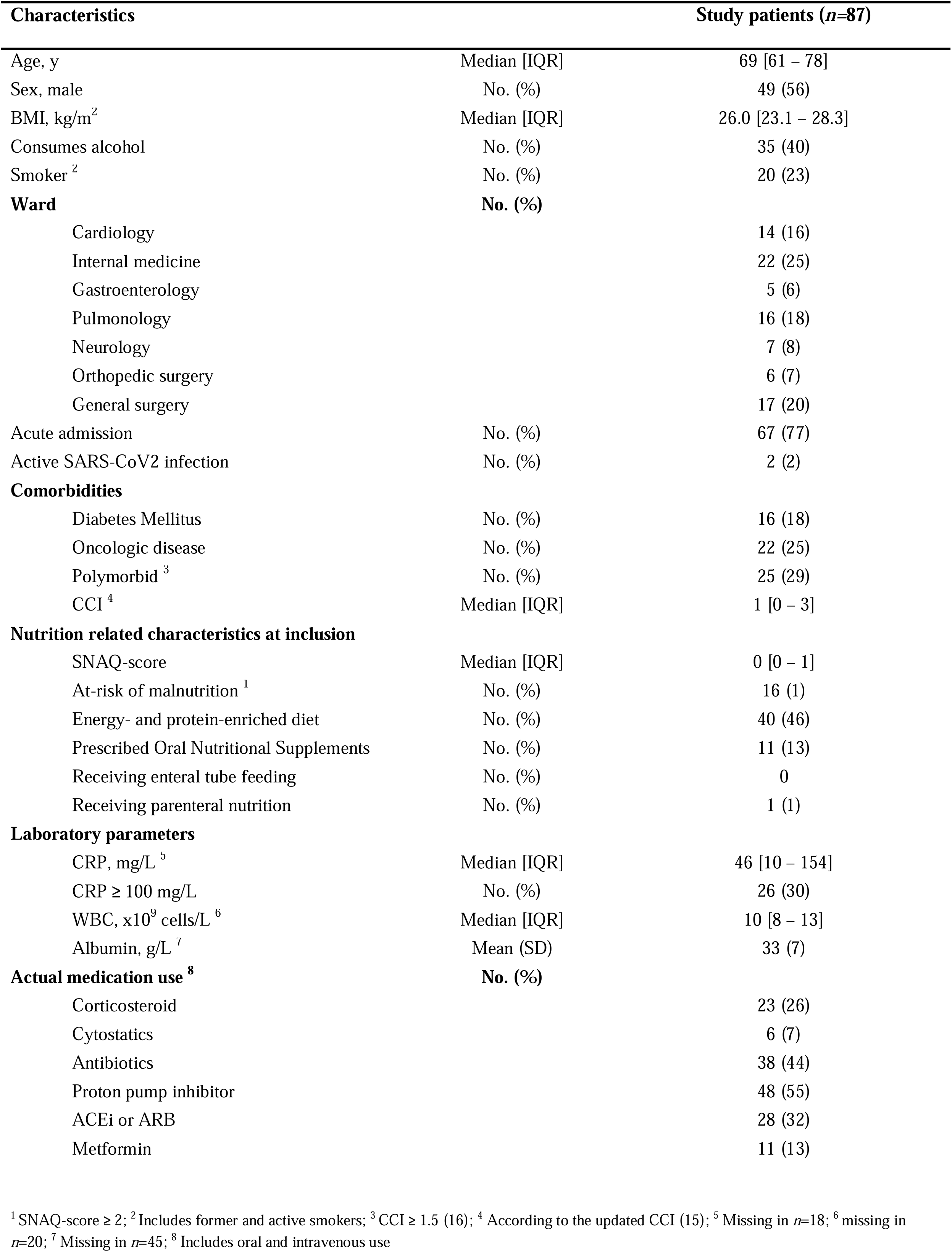
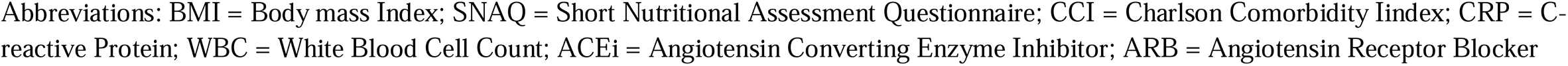
Characteristics of study patients who received a personalized nutrition product.

#### Sensory evaluation

Among the study patients, 137 PNPs were distributed. Questionnaires regarding consumption experience were missing in 11 due to no consumption, one due to the development of delirium, and one due to discharge home between product distribution and the questionnaire. Among the 124 available questionnaires, the mean score (scale 1 – 9) was 5.7 (**±**2.1) for overall liking, 5.8 (**±**2.1) for taste, 5.7 (**±**2.0) for aftertaste, 5.6 (**±**1.9) for texture, 6.2 (**±**1.7) for appearance, and 6.5 (**±**1.9) for freshness. Physical complaints were reported in 9 questionnaires from 9 study patients following consumption (uncomfortable sensation in the throat *n*=2, coughing *n*=2, gastro-intestinal discomfort *n*=2, swallowing difficulties *n*=2, and dizziness *n*=1). Overall liking and taste evaluation per PNP health profile are presented in Supplemental Figures 1 and 2 (Supplemental file 2).

#### Product consumption

Median consumption of the PNP was 51% [IQR 19 – 100] and 43g [IQR 13 – 93]. Of 11 distributions, no consumption occurred due to nausea (*n*=4), emotional situation (*n*=2), lack of appetite (*n*=2), diarrhea (*n*=1), swallowing problems (*n*=1), or unknown reasons (*n*=1). On 48 (35%) occasions, the PNP was completely consumed. Reasons for not consuming the whole product were given in 67 questionnaires, and included disliking the product in 58%, lack of appetite in 24%, disliking and lack of appetite in 3%, and not feeling well in 15%. Consumption rates (%) were correlated with overall liking, taste, texture, and freshness scores (R^2^ = 0.50, *p* < 0.001, R^2^ = 0.49, *p* < 0.001, R^2^ = 0.46, *p* < 0.001, and R^2^ = 0.37, *p* < 0.001, respectively), and product appearance scores (R^2^ = 0.22, *p* = 0.015). No differences in consumption rate were observed between the different PNP’s health profiles (*p* > 0.05; Supplemental Figures 3 and 4). Males consumed more than females (median 100% [IQR 33 – 100] vs. 38% [IQR 12 – 100], *p* = 0.021, and 48 g [IQR 27 – 72] vs. 25 g [IQR 8 – 55], *p* = 0.008, respectively). Serum albumin concentrations at hospital admission were correlated with the proportion of consumed product (R^2^ = 0.31, *p* = 0.043). In addition, age was negatively correlated with absolute consumption (R^2^ = -0.23, *p* = 0.030), but not with relative consumption (R^2^ = -0.10, *p* = 0.369). No correlations between other clinical characteristics (BMI, SNAQ-score, CRP levels, WBC, medication use) and product consumption (% and weight) were observed (Supplemental Tables 2 and 3, Supplemental file 2). The PNP provided a median of 71 kCal [IQR 24 – 137] and 1.4 g [IQR 0.4 – 3.4] protein. Among distributions of the PNP’s health profiles, muscle function (*n*=30) provided the highest median energy (151 kCal [IQR 54 – 305]) and protein (12.3g [IQR 4.4 – 25.9]) intake (Supplemental Figure 5 and 6, Supplemental file 2).

#### Reordering

The likeliness to reorder the PNP (scale 1 – 7) was 3.5 (**±**2.3) among 124 completed questionnaires. Among these, 83 times one or two reasons for not reordering were given, and 49 times a reason for reordering. 91% of the reasons for not reordering the PNP were due to sensory properties of the PNP, with the main reason not intending to reorder was finding the product “unpalatable” (52%). The main reason for intending to reorder the PNP was finding the product “tasteful” (78%). All reasons for reordering and not reordering the PNP are presented in Supplemental Figures 7 and 8 (Supplemental file 2), respectively. In 63% of the eligible moments to reorder a PNP for the following day (79 in total) a PNP was reordered, while the likeliness to reorder the product was scored 3.8 (±2.5) among these. Consumption rates and overall liking in study patients who reordered (*n*=34) compared to those who received only one distribution of the product (*n*= 53) did not differ; 50% [IQR 32 – 76] vs. 47% [IQR 15 – 100], *p* = 0.863, and 5.9 (±1.9) vs. 5.3 (±2.1), *p* = 0.193, respectively.

#### Attitude and perceptions

Among the 83 available questionnaires regarding attitude towards the concept, the value (scale 1 – 7) of a PNP service in a hospital setting was scored 5.3 (**±**1.7). The agreement (scale 1 – 7) that these PNPs may benefit patients’ physical and mental health was scored 5.3 (**±**1.8) and 4.8 (**±**1.7), respectively. The agreement (scale 1 – 7) of being worried about the use of personal information in a PNP service was scored 1.7 (**±**1.3). Despite incomplete disclosure regarding the production technique of the PNP, 22% of the participating patients reported that they were aware that their PNP was produced using 3D printing. Overall liking did not differ between those reported to be aware vs. unaware of the product being 3D-printed (5.8 ±1.7 vs. 5.4 ±2.2, *p* = 0.554). Patients who were aware of the 3D printing technology used to produce the PNP rated the service value higher than those who were unaware (5.9 ±0.9 vs. 5.0 ±1.9, *p* = 0.010).

### Staff

#### Sensory evaluation and acceptability

Fifteen staff members participated in the tasting and information session, consisting of dietitians *n=*7, chefs *n=*2, meal service center employees *n=*2, senior nurses *n=*2, and hospital ward managers *n=*2. One dietitian did not fill out the questionnaire. The PNP’s overall liking (scale 1 – 9) was 5.8 (**±**1.6). Taste, texture, appearance, and freshness of the PNP were scored 5.9 (**±**2.2), 5.5 (**±**2.0), 6.3 (**±**1.7), and 6.7 (**±**1.6), respectively. Agreement scores (1 – 7 scale) on attitude towards the concept, perceived benefits, and intentions to adopt and implement a PN service are presented in Table 2. Agreement on the statement ”“*I would like to know the benefits of these personalized food products on the health of hospitalized patients*” was scored highest with an average of 5.6 (±1.7).

**Table 2:**
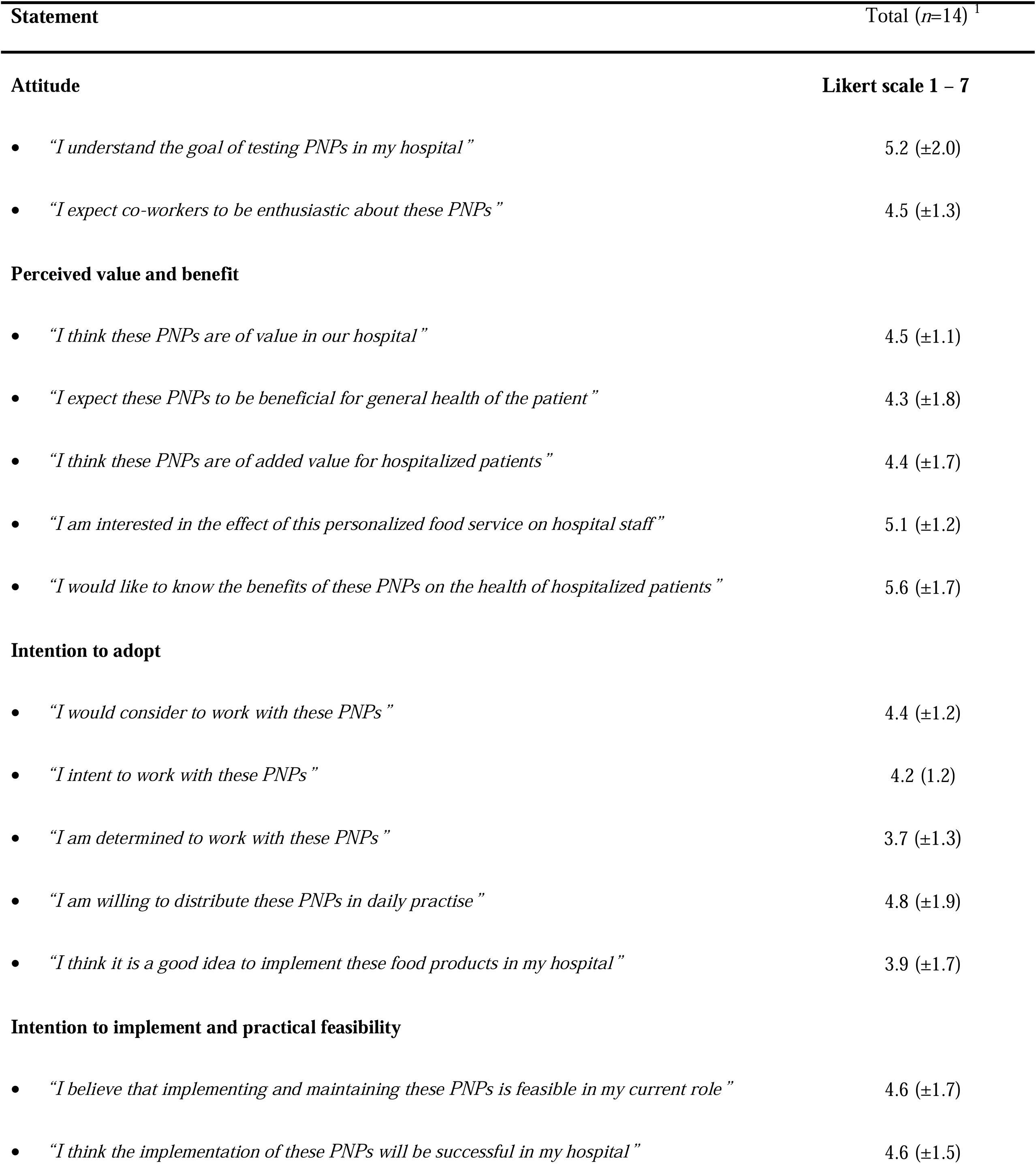
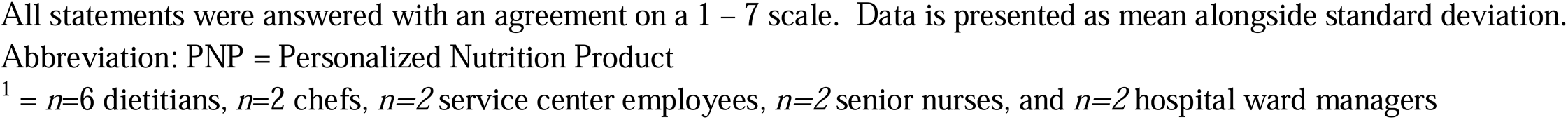
Agreement scores of hospital staff on attitude, perceived benefits, and intentions to adopt and implement a personalized nutritional product service.

## Discussion

To the best of our knowledge, this proof-of-concept study is the first to evaluate the experiences of a PNP service, including assessment of sensory liking, consumption rates, and attitudes among hospitalized patients and hospital staff. Sensory liking of the PNPs was moderately positive, with average overall liking scores of 5.7 on a 1 – 9 Likert scale, alongside PNP consumption rates of 51% among patients. Despite neutral intentions to reorder, actual reordering rates were 63% and a PNP service was scored as valuable by both patients and staff members. These findings provide novel insights into the acceptance of this innovative PN adjunct to conventional hospital nutrition.

### Sensory evaluation

Overall liking of the PNP was rated modestly positive by participating patients and staff with a mean of 5.7 and 5.8, respectively, on a 1 – 9 scale. In addition, product overall liking and taste scores appeared to be related to the product’s health profile, which differed in sizes, dough consistency, added micronutrient powders, and filling flavour options, similar to the observations among military subjects (Van Der Haar 2026 *Unpublished*). While no previous studies have investigated the sensory evaluation of fresh 3D-printed products in clinical settings, prior research in care homes identified a preference for cold and sweet snacks among patients (17), aligning with the type of PNPs in this study. Aside from sensory properties, product familiarity is shown as factor of great importance to enhance nutritional intake among hospitalized older patients (18). Here, we observed however no relation between age and overall liking or consumption rate. Prior surveys among cancer patients undergoing treatment revealed a preference for specific food properties such as fibre-rich dough and cheese or fruit fillings (19). Moreover, a qualitative study emphasized the importance of including savoury snack options in hospital settings (20), which were not optional in our PNP. We observed that taste and texture evaluation of the PNP were important determinants of partially consuming or reordering the product, highlighting the need of refinement of sensory attributes and flavour options to enhance patient adherence.

### Consumption rates

Consumption rates showed a wide distribution [IQR 19 – 100%] and 8% of the PNPs were not consumed. While product liking partly influenced consumption (R^2^ = 0.50), physiological symptoms that impact food intake such as the loss of appetite likely played a role. Prior research indicates that >50% of hospitalized patients consume only half or less of their ordered meals, and 58% skip in-between meal snacks (21). Factors such as older age, lower BMI, female sex, and acute admission have been associated with reduced meal consumption during hospitalization (6, 21). Moreover, systemic inflammation has been related to appetite suppression, which is thought to be a protective mechanism to prevent overfeeding-related autophagy (22). In our data, in addition to males consuming more than females, we observed a weak correlation between serum albumin levels, which is conversely associated to inflammation and PNP consumption rates (R^2^ = 0.31). These findings highlight the potential role of clinical factors and inflammatory biomarkers in influencing PNP consumption, underscoring the need for future studies to examine nutrition-impacting determinants to further optimize PN strategies.

Adherence to dietary interventions has been shown to be higher when nutrition is tailored to individual needs. In a large, multicenter, randomized controlled trial (RCT) on individualized nutritional support in medical in-patients at nutritional risk, patients who received individualized nutrition had better adherence to dietary protocols compared to those with no dietary consultation (1). Similarly, a recent RCT in healthy adults reported 30% higher adherence to personalized dietary advice than to general advice (23). The current study translated personalized nutritional advice directly into a personalised food product, which differs from previous PN interventions. Recently, it was shown that PN advice translated into personalised food boxes improved the ease of implementing nutritional guidance and decreases in body weight and hip circumference, compared with personalised nutrition advice or generic advice in adults living with overweight or obesity (24). Whether such personalised delivery formats, in the form of PNPs, similarly enhance long-term adherence and health outcomes in hospitalized patients remains to be investigated in future studies.

### Acceptability among patients

New food products often face challenges in gaining consumer acceptance due to their novelty and lack of familiarity among users (12). This may partly explain the neutral likeliness to reorder (mean 3.8 ±2.5 on 1 – 7 scale) observed in this study, as similar hesitancies could influence acceptance of PNPs. However, no differences in consumption rates between patients who reordered and those who declined reordering suggest that consumption rate was independent of the interest in PN services. In addition, patients rated the PN service as valuable (mean 5.3 ±1.7 on 1 – 7 scale), with minimal concerns about the use of personal information for customization. These are important findings as negative consumer attitudes towards the concept and privacy concerns have been suggested to limit the implementation of PN (25). Lastly, the use of novel production technologies, such as 3D food printing, have been suggested to hamper the acceptability of these products among consumers (12). While we applied incomplete disclosure regarding the production process of the PNP at inclusion of the study, one fifth was aware of the 3D food printing technique. These participants valued the PNP-service higher than those unaware, with comparable overall product liking between groups, suggesting that awareness of the technology’s role in product customization may enhance perceived benefits.

### Acceptability among hospital staff

Nutrition interventions may have the potential to improve in-patients’ food intake and nutritional status, but require acceptability and collaboration from multidisciplinary ward staff, including nurses, dietitians, and physicians to enhance involvement and success rate (26, 27). In addition to patient’s acceptance, the observations on the attitudes and intention to adopt among hospital staff provided valuable insights into the feasibility of a PNP service in clinical care. Surveyed staff rated the concept as slightly valuable (mean 4.5 ±1.1 on 1 – 7 scale) and would consider working with a PNP service, but interest in the effects on hospital staff and patient’s health benefit was also evident. Qualitative studies among nutrition experts and health care professionals indicated that 3D food printing may improve quality of life for those who require texture-modified food (28, 29). However, several barriers to acceptance have been highlighted by dysphagia patients, their supporters, and health care professionals (29, 30), including concerns on the appearance of 3D printed foods and their ability to closely resemble traditional food products, which play a critical role in food satisfaction (29, 30). In our study, the appearance of the PNP scored relatively high (6.2 **±**1.7 on a scale of 1-9), and concerns on the use of personal information in a PNP service was low. Moreover, we observed, although in a small sample size, that the prevalence of reported symptoms following PNP consumption were low and were unlikely attributable to the product. Our findings may help to address concerns regarding user acceptance and introduce the concept of 3D-printed PNP into the field of clinical nutrition. Nevertheless, the efficacy of a PNP in a medical setting remains unknown and appears critical to enhance its acceptability and compliance among patients and hospital staff.

### Limitations and future perspectives

While this proof-of-concept study provides novel and comprehensive insights into the acceptability of a PNP service in a clinical environment, several limitations should be acknowledged. First, the small sample size, short study duration, and single-center design limits the generalizability of findings. However, this was a first pilot study and the prototype FFMP only allowed for limited production duration and amount of PNPs. In addition, lack of a control group or product hampers robustness of findings. Furthermore, specific patient groups, such as those with swallowing difficulties and cognitive impairment were excluded, limiting applicability of the findings in more vulnerable groups (i.e. geriatric or neurologic patients and critical illness survivors) who might particularly benefit from a (texture-modified) PN intervention (29). Future research should focus on the feasibility of a FFMP in diverse clinical settings, such as hospitals, nursing homes, and rehabilitation centers. This strategy includes integrating the production and distribution processes into routine clinical workflows to facilitate an on-demand PNP service, which was unfeasible in this study due to its short period. Advanced personalization methods, such as body composition analysis and metabolomics, may be incorporated to optimize personalization of product composition for the individual patient. Ultimately, research evaluating the cost-effectiveness and patient adherence of PNPs compared to conventional clinical nutritional products is warranted.

## Conclusions

The present study showed that both patients and staff evaluated PNPs in hospitalised patients as moderately positive and valuable. However, considering the platform’s prototype status, further optimization and personalization of the products is necessary before investigating clinical efficacy. Our findings pave the way for further exploration of innovative nutritional strategies, including PN with 3D food printing techniques, to enhance nutritional care in hospital settings.

## Data Availability

All data produced in the present study are available upon reasonable request to the authors

## Acknowledgements

The authors would like to express their gratitude to José van Uden for her coordination and supervision on the project, to Roland Zegers, Dolf Klomp, Peter Giesen, and Martyasa Putra for engineering and developing the FFMP, to Eugene van Someren for developing the digital ordering system for the PNP, and to Lotte Peters and Hannah Eggink for their support in developing and formulating the health nutritional profiles (TNO, the Netherlands). Furthermore, we would like to thank Bei Tan, Seyed-Ali Ghoreishy, and Jolanda Henket (Wageningen University and Research, Wageningen, the Netherlands) for developing and preparing food materials. We are also grateful to Anna Keetels and Sascha Grevelingen for their assistance in distributing the PNPs and collecting questionnaires from patients (Dutch Nutrition & Healthcare Alliance, Ede, The Netherlands). Lastly, we would like to thank all hospital patients and staff who participated in this study. Their time, cooperation, and valuable contributions made this research possible.

## Funding statement

This investigator-initiated study was part of a larger project with the aim to develop and test an FFMP for the production of personalized 3D-printed nutritional products. This project was part of a public-private partnership ‘IMAGINE: Innovations for the manufacturing and marketing of fully personalized food products’, financed by Topsector Lifesciences & Health and Topsector High Tech Systems & Materials, the Netherlands (grant number 21074 (WFBR) and 100340397 (TNO)) and co-funded by Ministry of Defence, GEA, Tate & Lyle, General Mills, Nissin, and SoliPharma. The funders were not involved in the study design, collection, analysis, interpretation of data, the writing of this article, or the decision to submit it for publication.

## Conflict of interest

ARHvZ reported receiving honoraria for advisory board meetings, lectures, research, and travel expenses from Danone-Nutricia, Rousselot, Baxter, GE Healthcare, Nestlé and Abbot. IWKK holds a Dutch Research Council Veni Fellowship (2024-2028) for a research program focused on improving nutrition support in (post)-ICU patients. The other authors have nothing to declare in relation to this study.

## Declaration of generative AI and AI-assisted technologies

During the preparation of this work, the author(s) did not use generative AI or any AI-assisted technologies.

## Author contributions

Conceptualization: MaxM, SvdH, FPMH, MU, MH, AR, MenrikeM, MWJN, ARHvZ, IWKK

Recruitment of subjects: MaxM, SvB, IWKK

Data curation: MaxM, SvdH, SvB, MU, KvB

Formal analysis: MaxM, KvB

Writing - Original Draft: MaxM, SvdH, FMPH, IWKK

Writing – Review and editing: MaxM, SvdH, FPMH, SW, SvB, MU, KvB, MH, AR, MenrikeM, MWJN, ARHvZ, IWKK

Project administration: MaxM, MenrikeM, MWJN, IWKK

Methodology: MaxM, SvdH, FPMH, SW, MU, MenrikeM, ARHvZ, IWKK

Supervision: IWKK

## Notes

### Author Declarations

Ethics committee of Hospital Gelderse Vallei, Ede, the Netherlands gave ethical approval for this work

