## Supplemental Tables and Figures for "Personalized nutrition products in a hospital setting: A prospective proof-of-concept study of acceptability in hospitalized patients and staff"

### Supplemental Table 1a: Reasoning for macro- and micronutrient composition in PNP per health benefit

| **Health benefit** | **Dough composition** | **Reasoning for dough** | **Added amino acids or micronutrients** | **Reasoning for micronutrients** |
| --- | --- | --- | --- | --- |
| **Muscle health** | Protein and 25% reduced added free sugars | Higher protein content and use of high quality protein source to support muscle maintenance | Leucine, calcium | EFSA health claim on calcium to contribute to muscle functioning (1). Meta-analyses on increased protein and leucine content to be beneficial for muscle maintenance in older individuals and people with sarcopenia (2, 3) |
| **Immune health** | Cereal | High in dietary fibers to support immune health (4) | Zinc, vitamin C, folic acid | EFSA health claims on zinc, vitamin C, and folic acid to support the immune system (5-7). |
| **Wellbeing** | Carrot | Complex carbohydrates and dietary fibers to support mental health (8) | Folic acid | EFSA health claims on folic acid to reduce tiredness and fatigue and to support psychological function (9). |
| **Cognition** | Cereal, almond | Complex carbohydrates (10) and the nutrient profile of nuts, including almonds (11), may support cognitive function. | Iron, vitamin B12 | EFSA health claims for iron and vitamin B12 to contribute to cognitive function (12, 13) |
| **Energy restricted** | Protein (50% reduced added free sugars) | High protein content and low sugar content to stimulate satiety, low energy content for energy restriction (14). | Leucine | EFSA health claim on protein contributing to an increase in satiety leading to a reduction in energy intake (15). Leucine rich protein sources support satiety while preserving muscle mass (16). |
| **Blood glucose regulation** | Plum and no added free sugars | Low in free sugars and complex carbohydrates to reduce peaks in blood glucose levels (17) and improve glycaemic control (18). | N/A | N/A |

12.	EFSA Panel on Dietetic Products NaAN. Scientific Opinion on the substantiation of health claims related to iron and formation of red blood cells and haemoglobin (ID 374, 2889), oxygen transport (ID 255), contribution to normal energy-yielding metabolism

(ID 255), reduction of tiredness and fatigue (ID 255, 374, 2889), biotransformation of xenobiotic substances (ID 258), and “activity of heart, liver and muscles” (ID 397) pursuant to Article 13(1) of Regulation (EC) No 1924/2006. EFSA Journal. 2010;8(10):1740.

| **Health benefit** | **Dough** | **Estimated weight, g** | **Filling options** | **Energy, Kcal/100g** | **Carbohydrate, g/100g** | **Protein, g/100g** | **Fiber, g/100g** | **Amino acid or micronutrient enrichment (mg/pp)** |
| --- | --- | --- | --- | --- | --- | --- | --- | --- |
| **Muscle health** | Protein and 25% reduced added free sugars | 100 g | Ganache | 356 | 41.7 | 22.9 | 5.0 | Leucine ^1^ (825), calcium (1792) |
|  |  |  | Yoghurt/cream cheese/custard | 271 | 37.6 | 23.3 | 5.9 |  |
| **Immune health** | Cereal | 275 g | Apple | 204 | 28.3 | 2.8 | 5.0 | Zinc (52), vitamin C (196), folic acid (48) |
|  |  |  | Pear | 219 | 32.0 | 2.6 | 4.9 |  |
|  |  |  | Banana-strawberry | 187 | 23.7 | 2.7 | 4.3 |  |
| **Wellbeing** | Carrot | 75 g | Yoghurt/cream cheese/custard | 184 | 39.5 | 1.8 | 5.9 | Folic acid (48) |
| **Cognition** | Cereal, almond | 75 g | Ganache | 307 | 37.9 | 5.1 | 3.0 | Iron (226), vitamin B12 (51) |
|  |  |  | Pear | 218 | 31.7 | 4.2 | 2.5 |  |
| **Energy restricted** | Protein and 50% reduced added free sugars | 50 g | Banana-strawberry | 127 | 14.5 | 14.6 | 2.9 | Leucine^2^ (825) |
|  |  |  | Yoghurt/cream cheese/custard | 185 | 25.9 | 15.7 | 4.8 |  |
| **Blood glucose regulation** | Plum and no added free sugars | 50 g | Apple | 72 | 17.6 | 0.3 | 5.9 | NA |
|  |  |  | Banana-strawberry | 60 | 14.6 | 0.3 | 5.4 |  |
|  |  |  | Yoghurt/cream cheese/custard | 98 | 22.2 | 1.0 | 6.6 |  |

### Supplemental Table 1b: Composition of the PNPs per health benefit and filling

^1^ a casing naturally contains 2045mg leucine per product; ^2^ a casing naturally contains 1023mg leucine per product

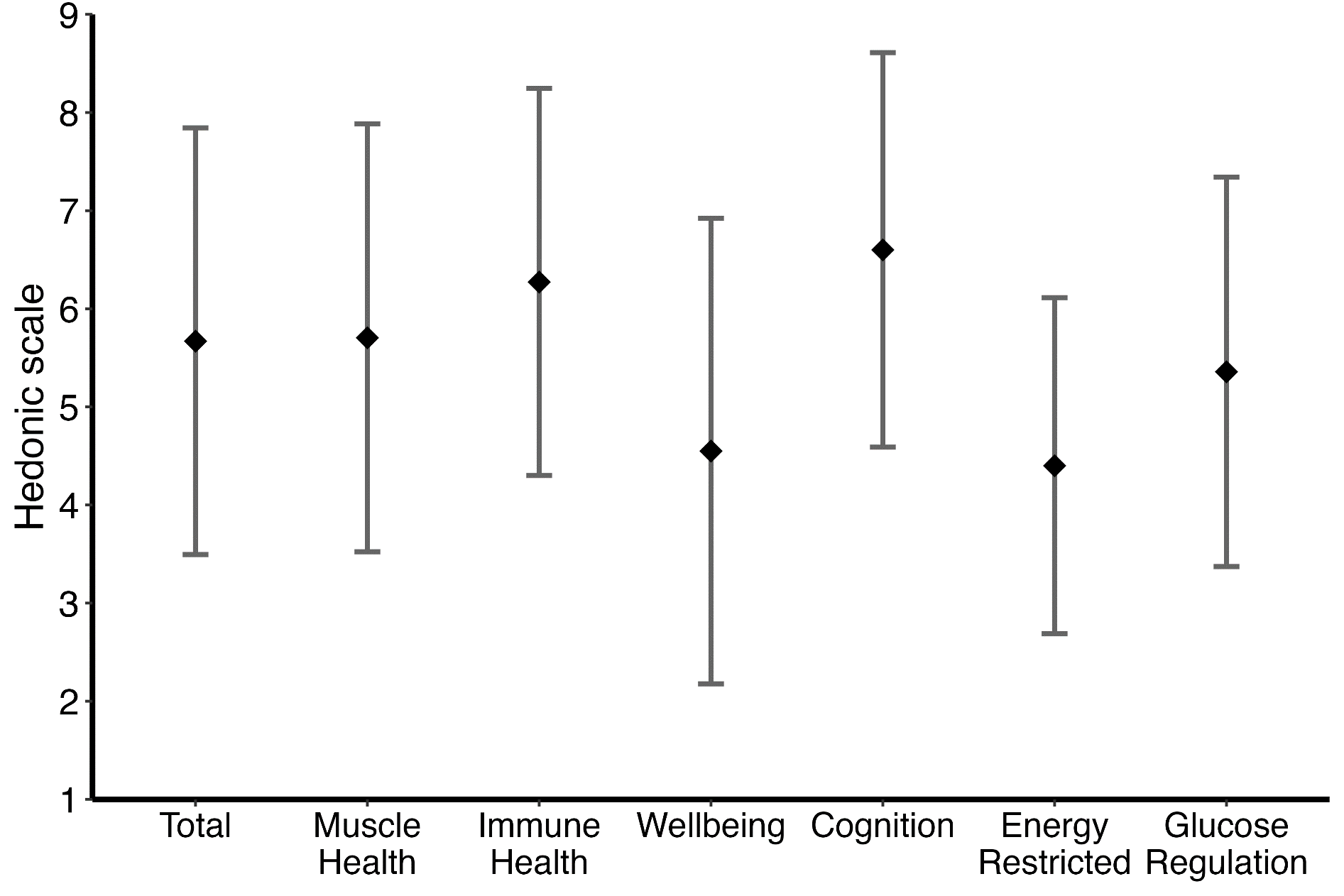

### Supplemental Figure 1: Overall liking per PNP’s health benefit

Means alongside standard deviation are displayed
Sensory evaluated PNPs: Muscle health, *n*=26; Immune health, *n=*33; Wellbeing, *n*=20; Cognition, *n*=21; Energy restricted, *n*=10; Glucose regulation, *n*=14

A significant difference in overall liking was observed between Cognition vs. Wellbeing (Student’s T-test, *p* = 0.004) and Cognition vs. Energy Restricted (*p* = 0.009). In addition, there was a significant difference between Immune health vs. Wellbeing (*p* = 0.007) and Immune Health vs. Energy Restricted (*p* = 0.013). No other pairwise comparisons reached statistical significance (all, *p* > 0.05).

### Supplemental Figure 2: Taste scores per PNP’s health benefit

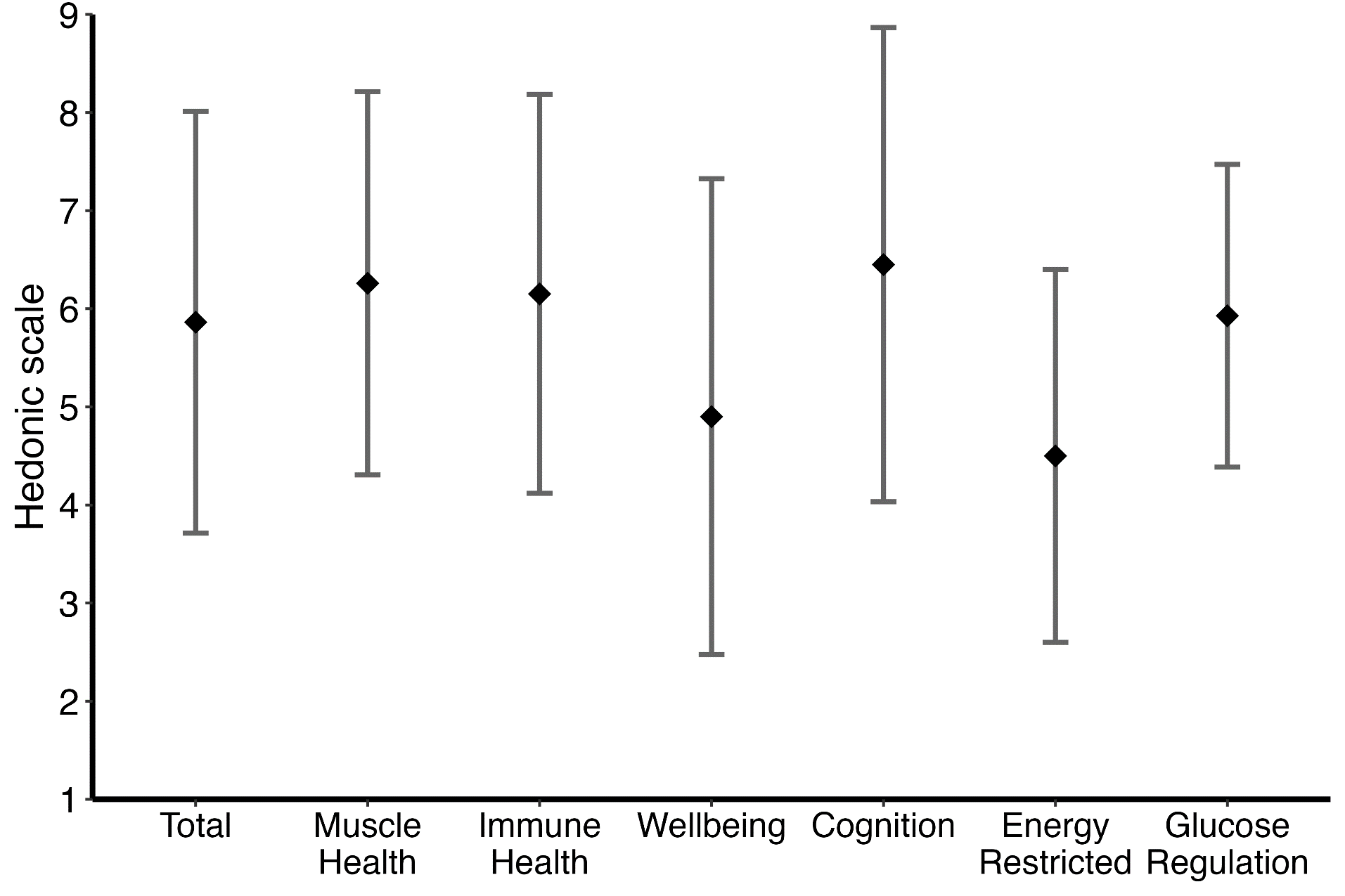

Means alongside standard deviation are displayed
Sensory evaluated PNPs: Muscle health, *n*=26; Immune health, *n=*33; Wellbeing, *n*=20; Cognition, *n*=21; Energy restricted, *n*=10; Glucose regulation, *n*=14
A significant difference in taste was observed between Cognition vs. Wellbeing (*p* = 0.046) and Cognition vs. Energy Restricted (*p* = 0.022). In addition, there was a significant difference between Immune Health vs. Energy Restricted (*p* = 0.027) and Muscle Health vs. Energy Restricted (*p* = 0.034). No other pairwise comparisons reached statistical significance (all, *p* > 0.05).

### Supplemental figure 3: Consumption (%) per PNP’s health benefit

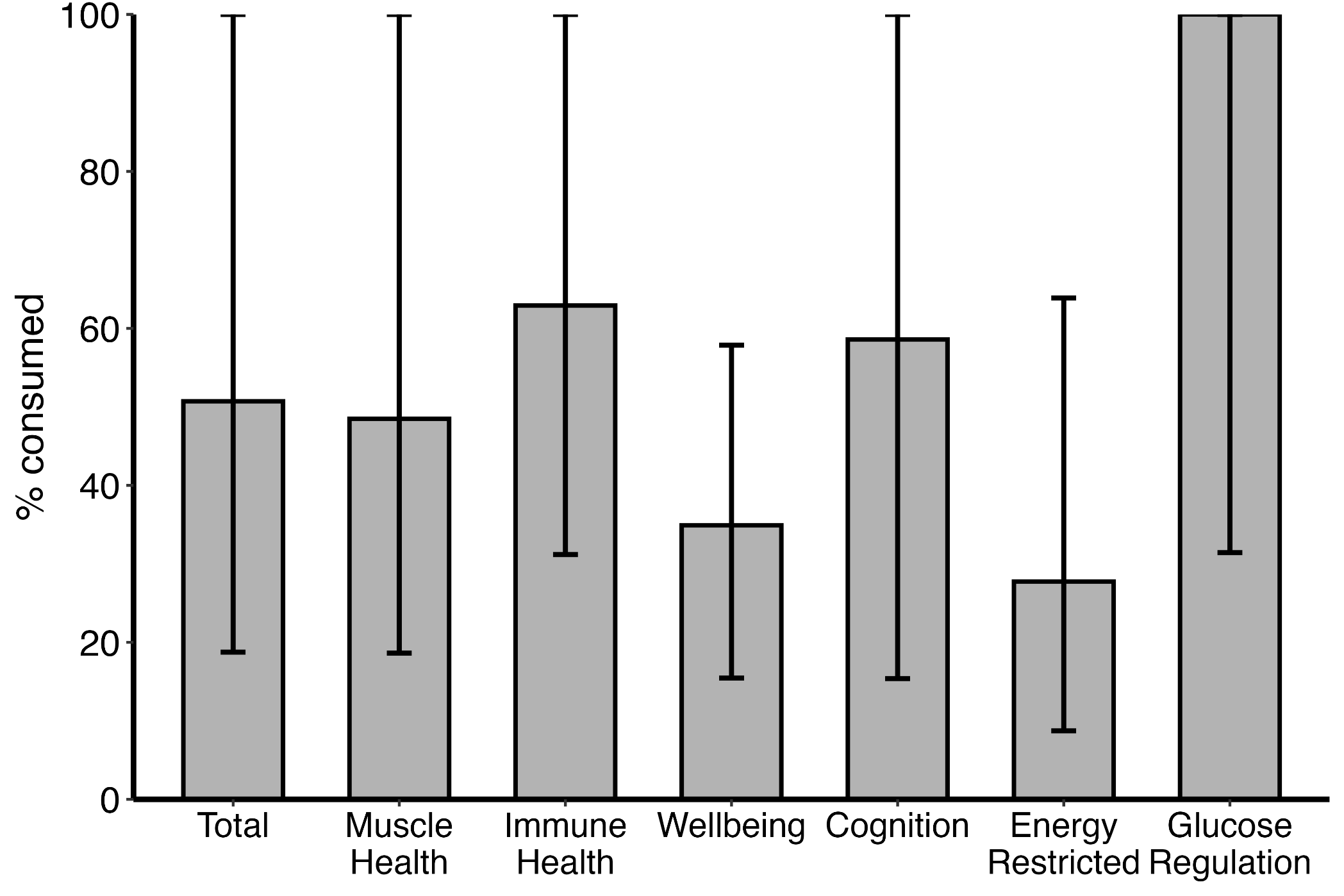

Medians alongside interquartile ranges are presented.

Distributed PNPs: Muscle health, *n*=30; Immune health, *n=*38; Wellbeing, *n*=21; Cognition, *n*=23; Energy restricted, *n*=11; Glucose regulation, *n*=14

No significant differences were observed in proportion consumed between health profiles (all, *p* > 0.05 using Mann Whitney U Test)

### Supplemental Figure 4: Consumption (g) per PNP health benefit

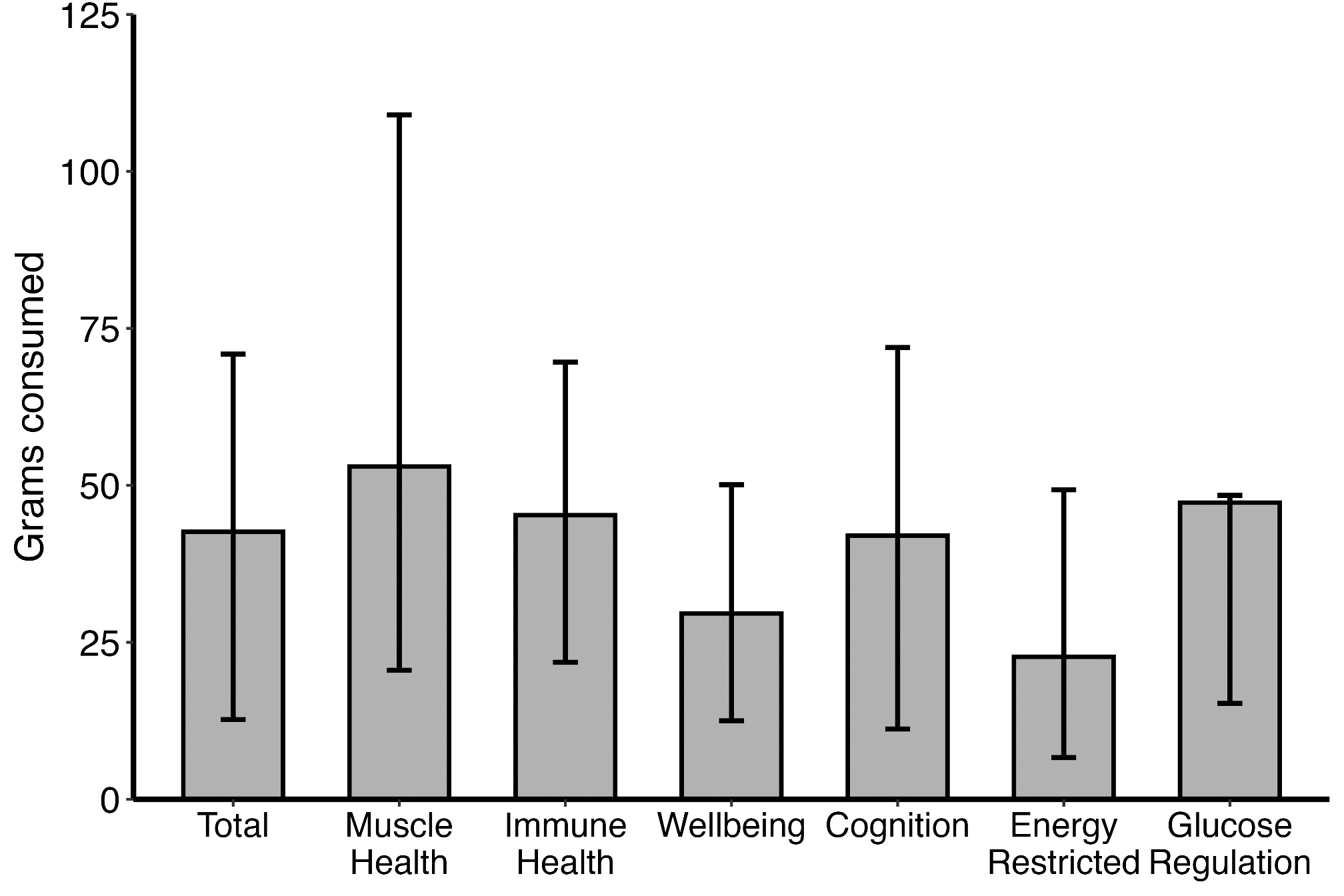

Medians alongside interquartile ranges are presented.
Distributed PNPs: Muscle health, *n*=30; Immune health, *n=*38; Wellbeing, *n*=21; Cognition, *n*=23; Energy restricted, *n*=11; Glucose regulation, *n*=14

No significant differences were observed in weight consumed between health profiles (all, *p* > 0.05 using Mann Whitney U Test)

| **Characteristics** | Consumption, % | *p*-value | Consumption, g | *p*-value | Overall liking score | *p*-value | Taste score | *p-value* |
| --- | --- | --- | --- | --- | --- | --- | --- | --- |
| **Male vs.**  **female** | 100 [33 – 100] vs.  38 [12 – 100] | 0.021* | 48 [27 – 72] vs.  25 [8 – 55] | 0.011* | 5.9 (**±**1.9) vs.  5.3 (**±**2.4) | 0.196 | 6.1 (**±**1.9) vs.  5.6 (**±**2.4) | 0.373 |
| **Obese vs.**  **non-obese** | 61 [22 – 100] vs.  66 [22 – 100] | 0.855 | 36 [7 – 74] vs.  46 [17 – 70] | 0.697 | 4.9 (±2.0) vs.  5.8 (±2.1) | 0.114 | 4.8 (±2.3) vs.  6.1 (±2.0) | 0.055 |
| **Acute admission vs. no acute admission** | 58 [19 – 100] vs.  100 [29 – 100] | 0.244 | 45 [12 – 70] vs.  49 [22 – 71] | 0.548 | 5.7 (± 2.2) vs.  5.5 (± 2.1) | 0.630 | 6.0 (± 2.1) vs.  5.5 (2.4) | 0.365 |
| **Risk of malnutrition vs.**  **no risk of malnutrition** | 53 [23 – 94] vs.  72 [21 – 100] | 0.608 | 40 [17 – 59] vs.  48[13 – 72] | 0.466 | 5.9 (±2.6) vs.  5.6 (±2.0) | 0.550 | 6.2 (±2.5) vs.  5.8 (±2.0) | 0.525 |
| **Polymorbid vs.**  **non-polymorbid** | 56 [24 – 100] vs.  75 [20 – 100] | 0.866 | 46 [15 – 55] vs.  48 [12 – 72] | 0.569 | 5.8 (±2.2) vs.  5.5 (±2.1) | 0.539 | 6.0 (±2.2) vs.  5.7 (±2.1) | 0.553 |
| **Smoker vs.**  **non-smoker** | 86 [27 – 100] vs.  58 [21 – 100] | 0.557 | 48 [15 – 63] vs.  45 [13 – 71] | 0.856 | 5.7 (±2.0) vs.  5.6 (±2.2) | 0.841 | 5.7 (±2.2) vs.  5.9 (±2.1) | 0.743 |
| **Alcohol consumer**  **vs. non-consumer** | 100 [25 – 100] vs.  47 [19 – 100] | 0.174 | 53 [23 – 73] vs.  38 [13 – 56] | 0.078 | 5.5 (±2.0) vs.  5.8 (±2.2) | 0.593 | 5.7 (±2.0) vs.  6.0 (±2.2) | 0.612 |
| **Energy and protein enriched diet**  **vs. no enriched diet** | 47 [16 – 100] vs.  100 [25 – 100] | 0.218 | 34 [12 – 63] vs.  48 [21 – 72] | 0.188 | 5.6 (±2.3) vs.  5.7 (±1.9) | 0.807 | 5.9 (±2.2) vs.  5.8 (±2.1) | 0.943 |
| **ONS user**  **vs. non-user** | 66 [30 – 100] vs.  61 [21 – 100] | 0.811 | 46 [40 – 60] vs.  47 [12 – 71] | 0.730 | 5.8 (±2.3) vs.  5.6 (±2.1) | 0.780 | 6.6 (±2.4) vs.  5.8 (±2.1) | 0.270 |
| **CRP ≥ 100**  **vs. < 100 mg/L** | 46 [12 – 100] vs. 75 [27 – 100] | 0.102 | 36 [10 – 65] vs.  49 [23 – 73] | 0.070 | 5.6 (±2.3) vs. 6.0 (±2.0) | 0.545 | 5.7 (2.2) vs.  6.4 (1.8) | 0.185 |
| **Corticosteroid user**  **vs. non-user** | 47 [23 – 100] vs.  70 [19 – 100] | 0.818 | 32 [21 – 69] vs. 48 [12 – 78] | 0.985 | 5.3 (±2.1) vs.  5.8 (±2.1) | 0.360 | 5.5 (±2.1) vs.  6.0 (±2.2) | 0.293 |
| **Metformin user**  **vs. non-user** | 62 [22 – 100] vs.  100 [25 – 100] | 0.465 | 46 [13 – 48] vs.  47 [14 – 72] | 0.269 | 4.4 (±1.5) vs.  5.7 (±2.2) | 0.633 | 5.7 (±1.6) vs.  5.9 (±2.2) | 0.802 |
| **Cytostatic user**  **vs. non-user** | 79 [25 – 100] vs.  66 [22 – 100] | 0.676 | 52 [31 – 86] vs.  46 [13 – 70] | 0.451 | 6.0 (±2.0) vs. 5.6 (±2.1) | 0.678 | 5.8 (±2.1) vs.  5.8 (±2.2) | 0.957 |
| **Antibiotic user**  **vs. non-user** | 47 [12 – 100] vs.  72 [27 – 100] | 0.336 | 41 [11 – 69] vs.  48 [23 – 71] | 0.334 | 5.9 (±2.3) vs.  5.4 (±2.0) | 0.242 | 6.1 (±2.2) vs.  5.7 (±2.1) | 0.396 |
| **ACEi/ARB user**  **vs. non-user** | 43 [22 – 100] vs.  73 [19 – 100] | 0.520 | 29 [12 – 54] vs.  48 [17 – 72] | 0.313 | 4.9 (±1.9) vs.  6.0 (±2.1) | 0.022* | 5.2 (±1.8) vs.  6.2 (±2.2) | 0.051 |

### Supplemental Table 2: Consumption rates, taste, and overall liking scores among binary characteristics

Median alongside [IQR] are reported for consumption rates and compared between groups by Mann-Whitney U test. Taste and overall liking scores are reported as mean with (±SD) and between groups compared using Student’s T-test.
Abbreviations: ONS = Oral Nutritional Supplements; CRP = C-reactive Protein; ACEi = Angiotensin Converting Enzyme inhibitor; ARB = Angiotensin Receptor Blocker

* significant difference, *p*<0.05

| **Characteristics** | Consumption, % | *p*-value | Consumption, g | *p*-value | Overall liking score | *p*-value | Taste score | *p-value* |
| --- | --- | --- | --- | --- | --- | --- | --- | --- |
| **Age** | -0.098 | 0.369 | -0.233 | 0.030 | 0.045 | 0.686 | -0.08 | 0.944 |
| **BMI** | 0.082 | 0.450 | 0.035 | 0.746 | -0.120 | 0.280 | -0.280 | 0.100 |
| **CRP** | -0.234 | 0.053 | -0.236 | 0.051 | -0.030 | 0.815 | -0.177 | 0.355 |
| **WBC** | 0.115 | 0.354 | 0.111 | 0.371 | 0.129 | 0.314 | 0.051 | 0.692 |
| **Albumin** | 0.313 | 0.043 | 0.270 | 0.083 | 0.013 | 0.939 | 0.070 | 0.672 |
| **Overall liking score** | 0.495 | <0.001 | 0.449 | <0.001 | x |  | 0.815 | <0.001 |
| **Taste score** | 0.493 | <0.001 | 0.460 | <0.001 | 0.815 | <0.001 | x |  |
| **Texture score** | 0.461 | <0.001 | 0.415 | <0.001 | 0.686 | <0.001 | 0.607 | <0.001 |
| **Freshness score** | 0.370 | <0.001 | 0.350 | <0.001 | 0.622 | <0.001 | 0.588 | <0.001 |
| **Appearance score** | 0.218 | 0.015 | 0.228 | 0.011 | 0.408 | <0.001 | 0.396 | <0.001 |

### Supplemental Table 3: Correlations between continuous characteristics and consumption and sensory liking.

Pearson correlation coefficients are reported.

Abbreviations: BMI = Body Mass Index; CRP = C-reactive protein; WBC = White Blood Count

### Supplemental Figure 5: Protein consumption per PNP health profile

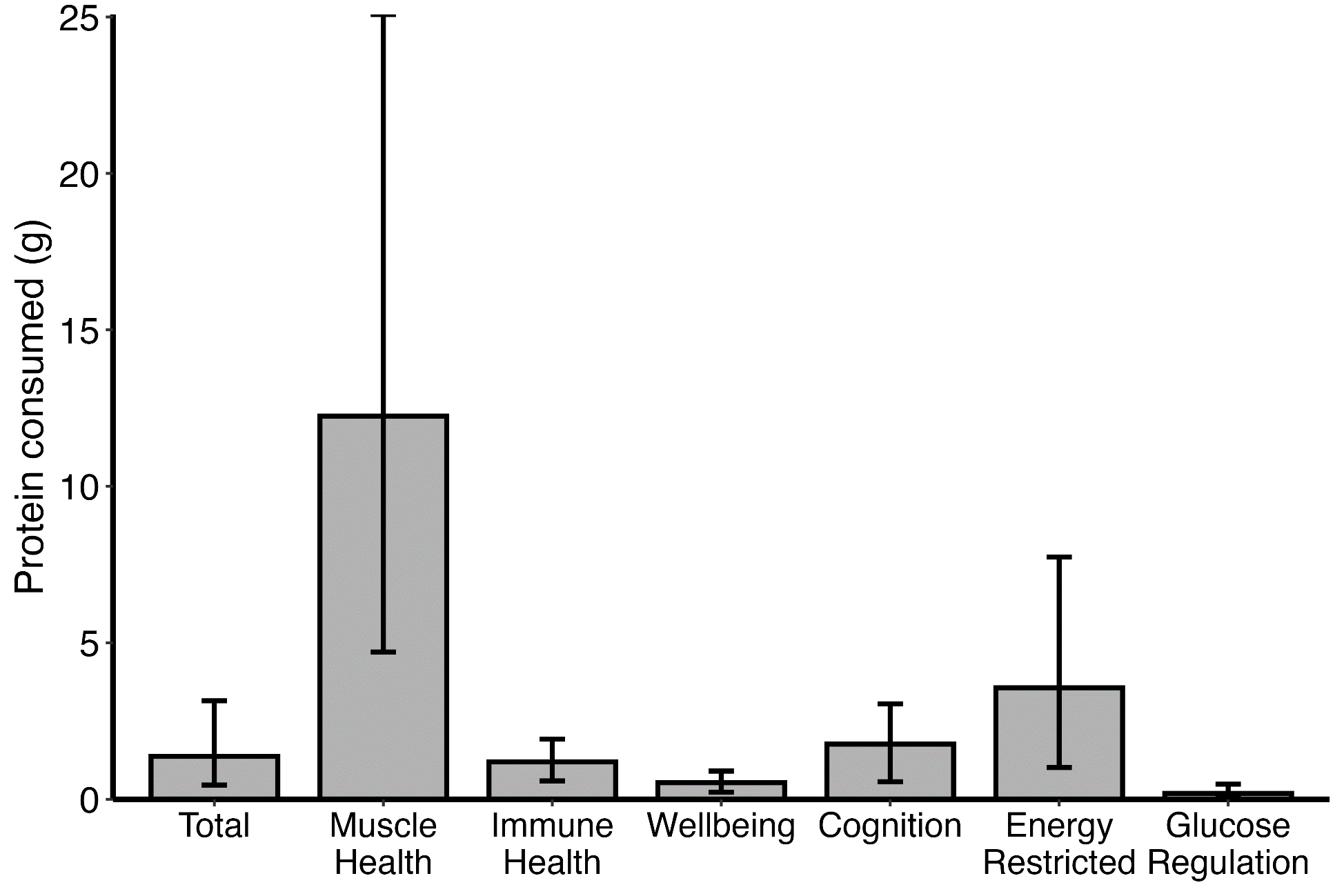

Medians alongside interquartile ranges are presented.
Distributed PNPs: Muscle health, *n*=30; Immune health, *n=*38; Wellbeing, *n*=21; Cognition, *n*=23; Energy restricted, *n*=11; Glucose regulation, *n*=14

### Supplemental Figure 6: Energy consumption per PNP health profile.

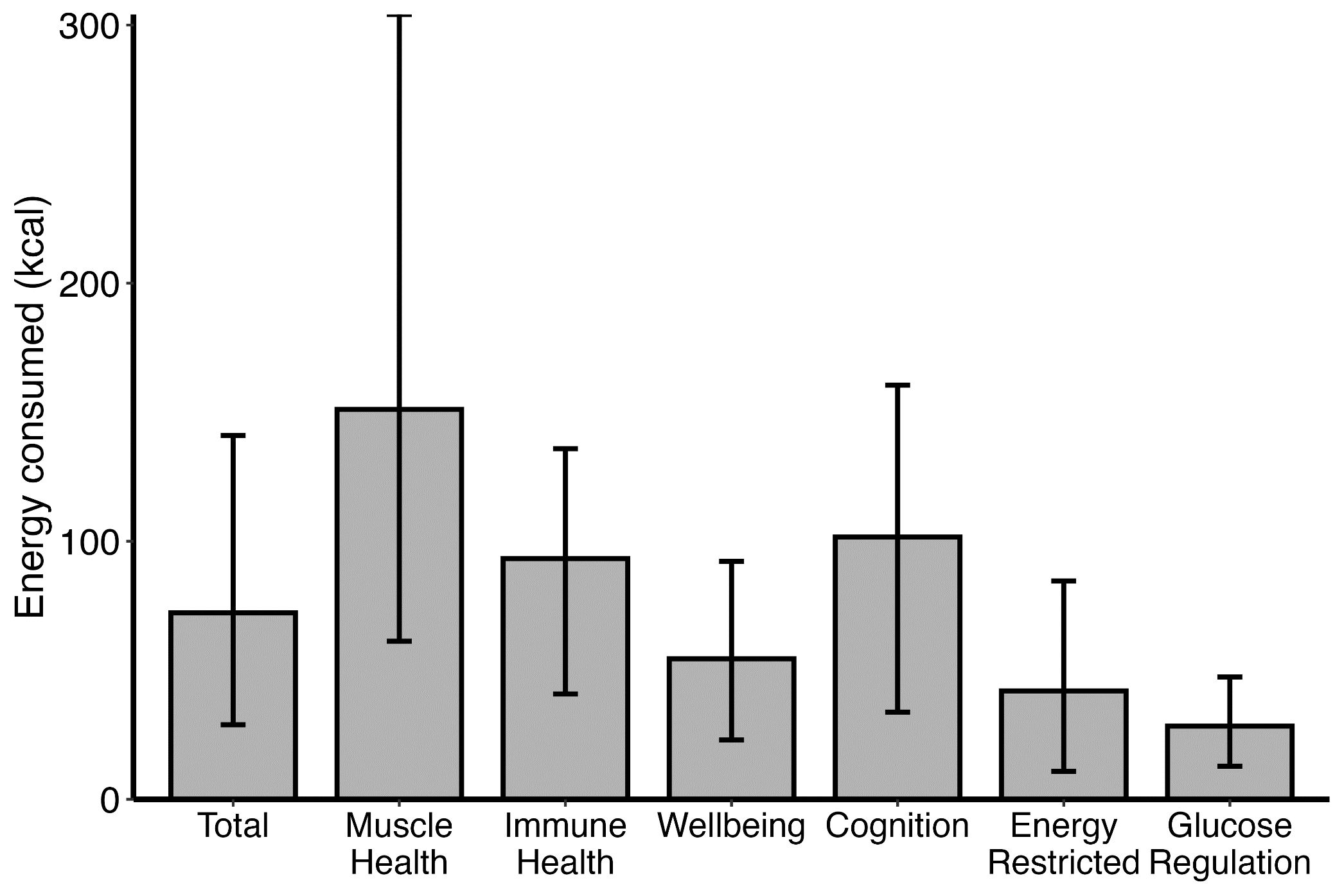

Medians alongside interquartile ranges are presented.
Distributed PNPs: Muscle health, *n*=30; Immune health, *n=*38; Wellbeing, *n*=21; Cognition, *n*=23; Energy restricted, *n*=11; Glucose regulation, *n*=14

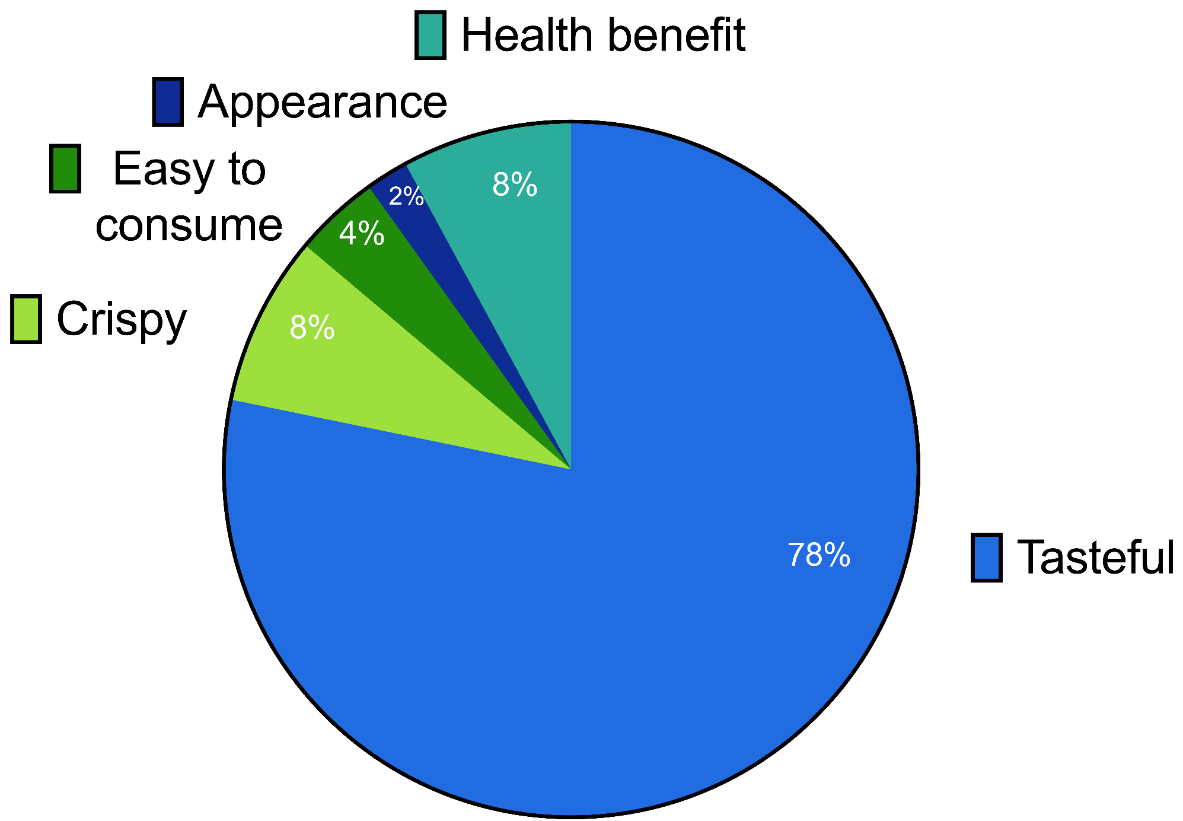

### Supplemental Figure 7: Reasons for intending to reorder a PNP

49 times one or multiple reasons for reordering were reported, of which once two reasons were reported.

### Supplemental Figure 8: Reasons for not intending to reorder a PNP

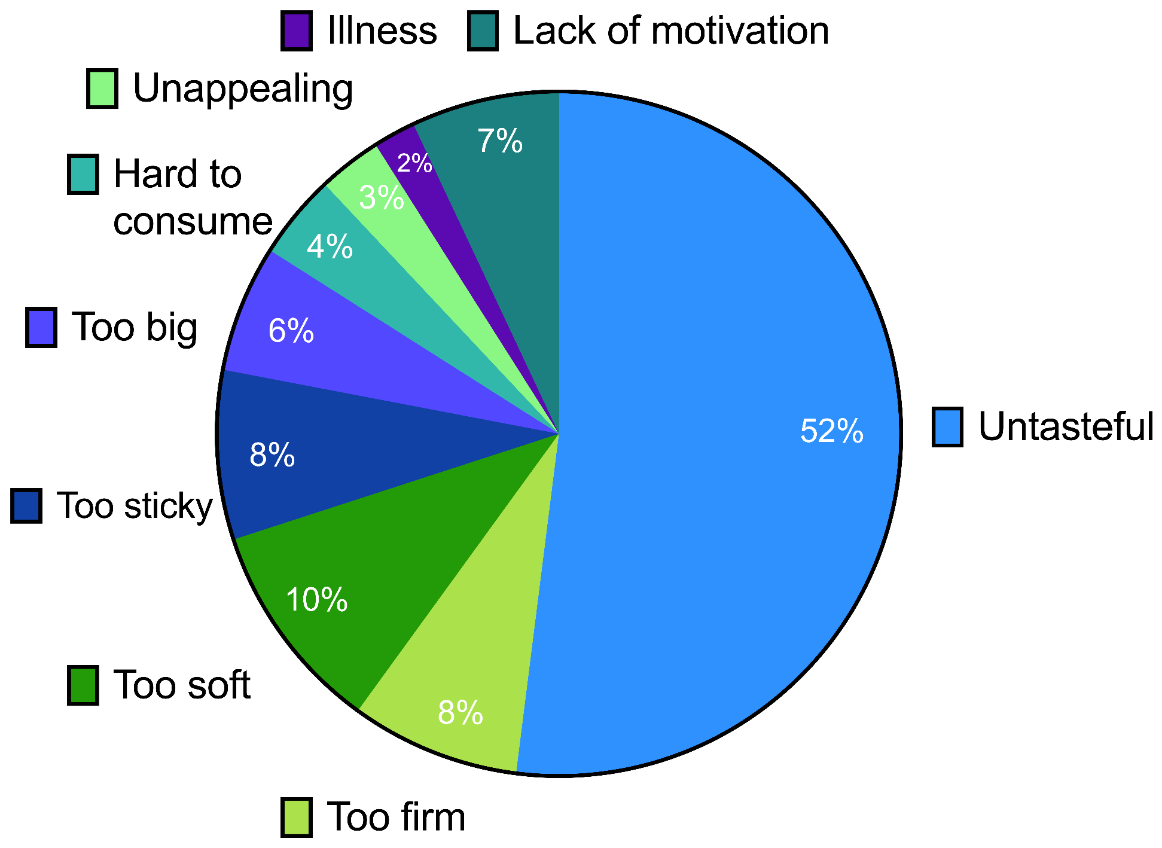

83 times one or multiple reasons for not reordering were reported, of which 7 times two reasons were reported.
