## Supplemental Methods for "Personalized nutrition products in a hospital setting: A prospective proof-of-concept study of acceptability in hospitalized patients and staff"

**Daily Questionnaire (Translated from Dutch to English)**

**Introduction**
Welcome to the daily questionnaire of the IMAGINE study. You are about to evaluate a NutriBite. This is a healthy and freshly prepared personalized nutrition product, enriched with vitamins and minerals, tailored to your personal preferences and needs. The following questions are about your experience with the NutriBite. You do not have to eat the entire NutriBite if you don’t want to. We are interested in your personal opinion, so there are no right or wrong answers.

**Questions about the product**

1.
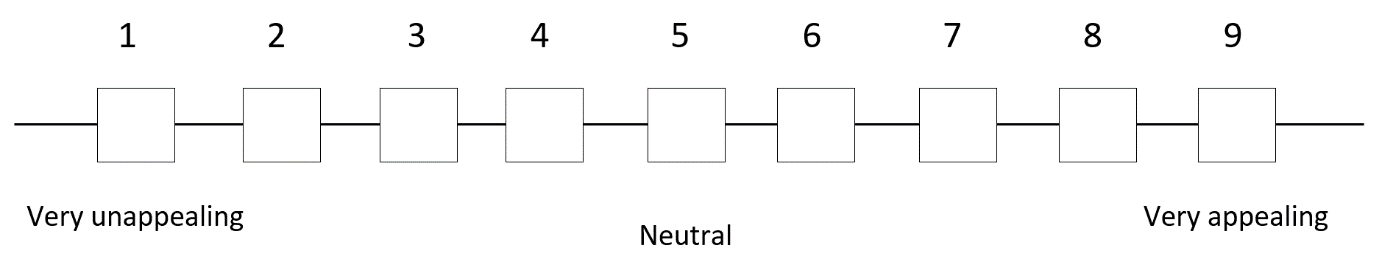
What do you think of the appearance of the NutriBite?
2. What do you think about the mouthfeel/texture of the NutriBite?

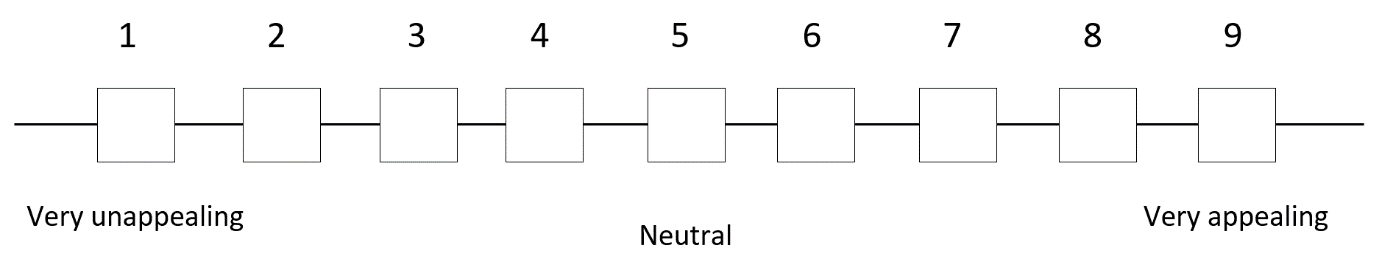

3.
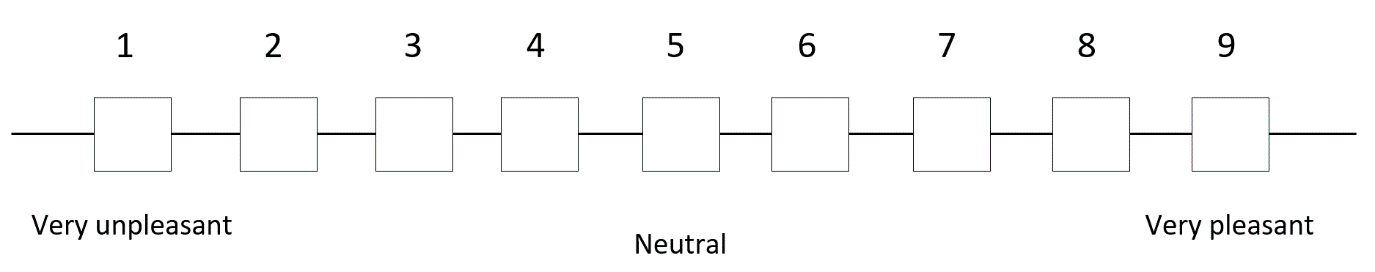
What do you think of the taste of the NutriBite?
4.
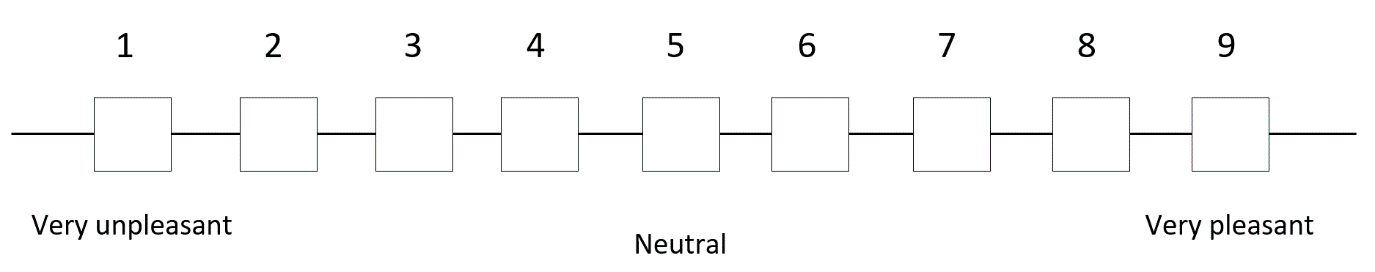
What do you think of the aftertaste of the NutriBite?
5. What do you think of the freshness of this NutriBite?

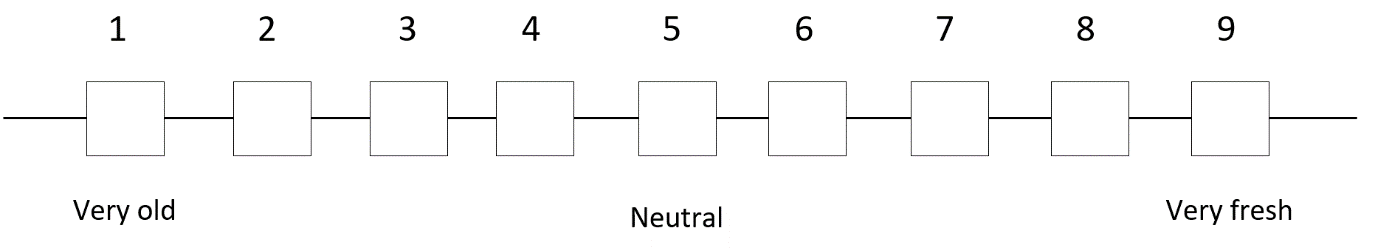

6.
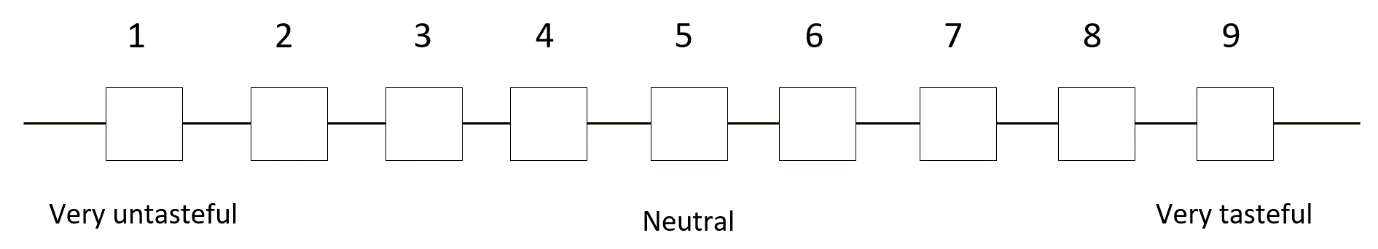
What do you think of this NutriBite overall (mouthfeel, taste and aftertaste)?
7. How can this NutriBite be improved?
8. Did you finish the NutriBite completely?

- Yes
- No

9. Why did you not finish the NutriBite completely?

- I did not like it
  - Why not? ________________________________________________________
- I was not hungry
- Other: _________________________________________________________________

10.
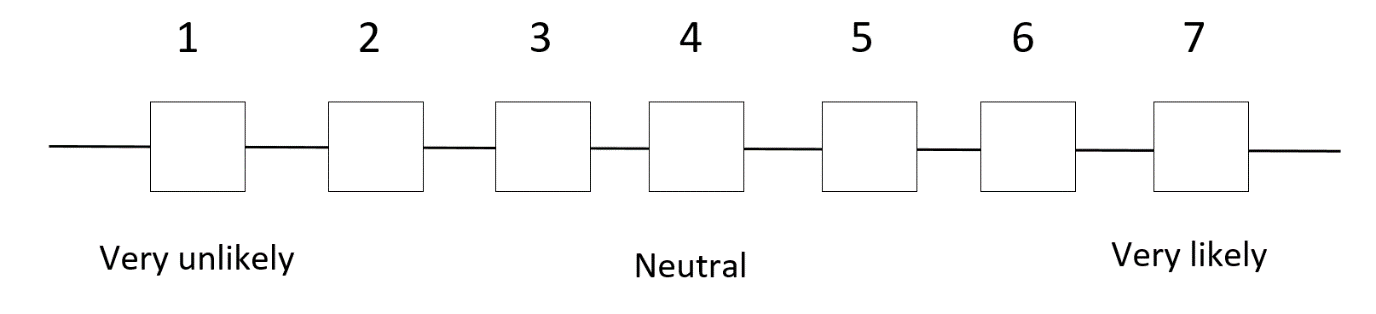
How likely would you re-order this NutriBite?
11. Why? ______________________________________________________________________
    a. Do you have any other remarks on the NutriBite?

________________________________________________________________________

12. Did you consume a NutriBite yesterday?

- Yes; continue to question 13
- No; continue with question 14

13. Did you experience anything after consumption that you would like to share?

___________________________________________________________________________

14. Would you like to order a NutriBite again for tomorrow? (*only if patient is deemed eligible by physician researcher to reorder)*

- Yes; continue to question 16
- No; continue to question 15

In case of no or in case the patient can not reorder again tomorrow due to logistic reasons:

15. The NutriBite was produced by a 3D food printer. Were you aware of this?

- Yes
- No

**Questions about a personalized nutritional service and -products**

Personalized nutrition products are tailored to your personal nutritional requirements and taste preferences. An example is a NutriBite you have just tasted. In the future it could be that a personalized nutrition service is implemented in the hospital. This operates by extracting your personal information (i.e. length, weight, medical information) and you provide taste preferences. Based on these information a fresh food product will be prepared that is tailored to your personal situation and potentially benefits your health.

16.
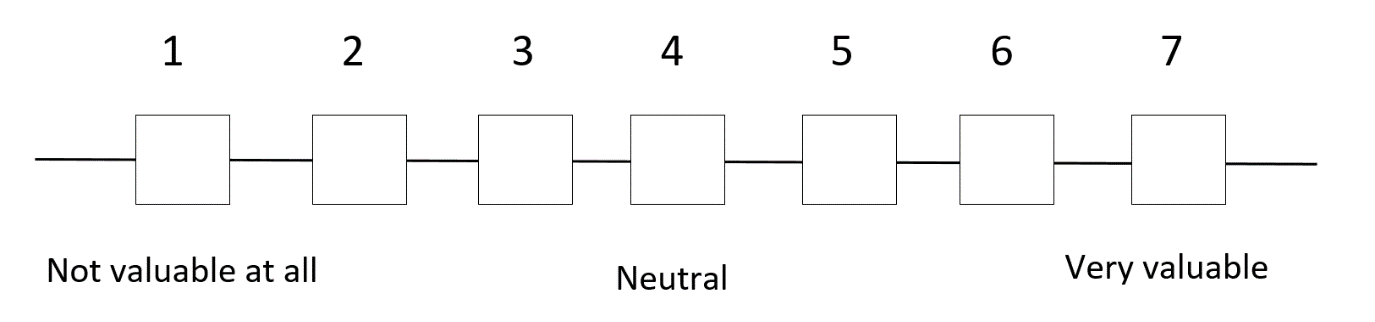
A personalized nutrition service in the hospital is….
17. I expect these products have a beneficial effect on my physical health

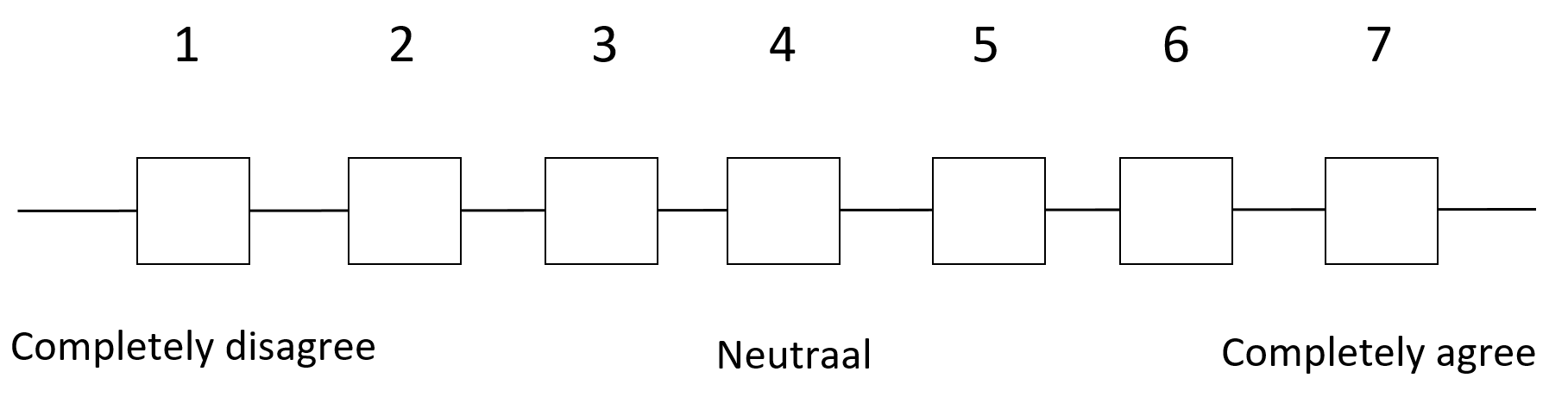

18. I expect these products have a beneficial effect on my mental health

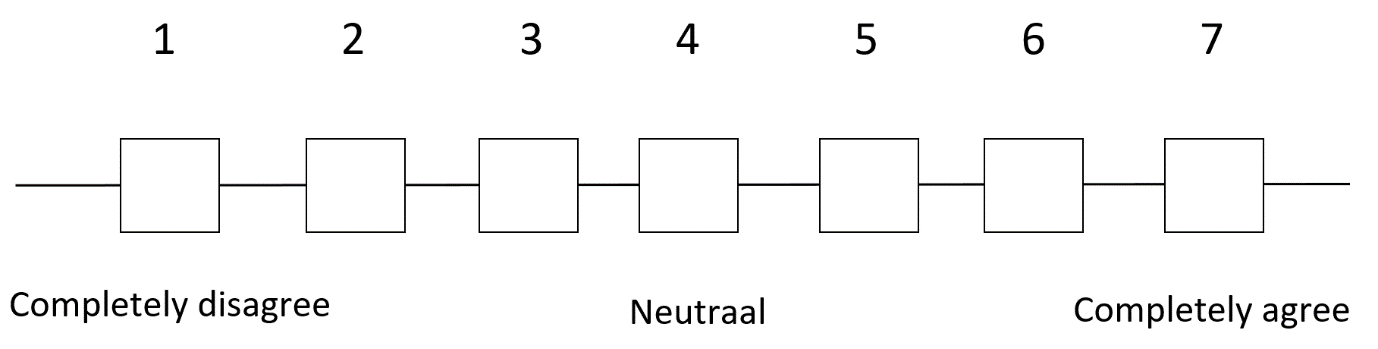

19. I find it concerning that my personal information is used in a digital personalized nutrition service system

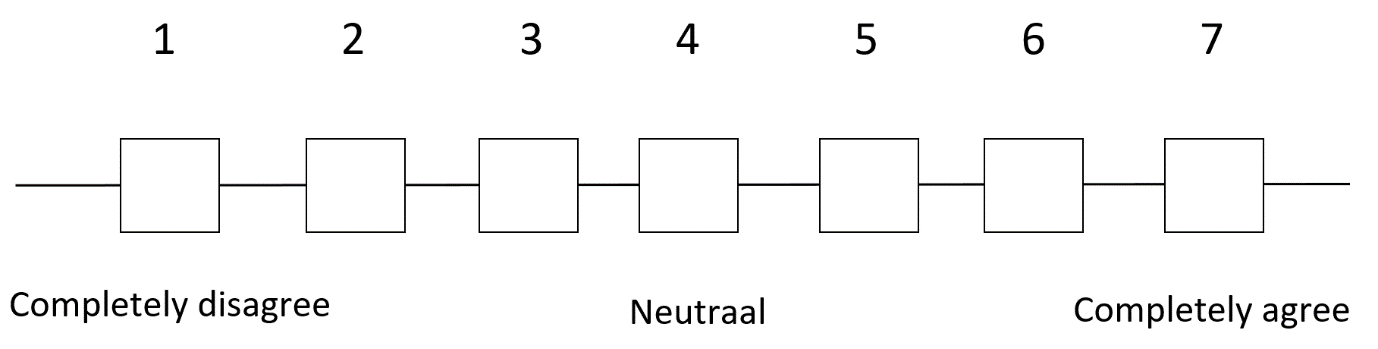

20. Do you anticipate any other advantages or disadvantages of a personalized nutrition service in the hospital?

- Yes; continue to question 21
- No;

21. What advantages or disadvantages do you expect?

_________________________________________________________________________________

End of questionnaire.
